# Cohort Profile: Post-hospitalisation COVID-19 study (PHOSP-COVID)

**DOI:** 10.1101/2023.05.08.23289442

**Authors:** Omer Elneima, Hamish J C McAuley, Olivia C Leavy, James D Chalmers, Alex Horsley, Ling-Pei Ho, Michael Marks, Krisnah Poinasamy, Betty Raman, Aarti Shikotra, Amisha Singapuri, Marco Sereno, Victoria C Harris, Linzy Houchen-Wolloff, Ruth M Saunders, Neil J Greening, Matthew Richardson, Jennifer K Quint, Andrew Briggs, Annemarie B Docherty, Steven Kerr, Ewen M Harrison, Nazir I Lone, Mathew Thorpe, Liam G Heaney, Keir E Lewis, Raminder Aul, Paul Beirne, Charlotte E Bolton, Jeremy S Brown, Gourab Choudhury, Nawar Diar Bakerly, Nicholas Easom, Carlos Echevarria, Jonathan Fuld, Nick Hart, John R Hurst, Mark G Jones, Dhruv Parekh, Paul Pfeffer, Najib M Rahman, Sarah L Rowland-Jones, AA Roger Thompson, Caroline Jolley, Ajay M Shah, Dan G Wootton, Trudie Chalder, Melanie J Davies, Anthony De Soyza, John R Geddes, William Greenhalf, Simon Heller, Luke S Howard, Joseph Jacob, R Gisli Jenkins, Janet M Lord, William D-C Man, Gerry P McCann, Stefan Neubauer, Peter JM Openshaw, Joanna C Porter, Matthew J Rowland, Janet T Scott, Malcolm G Semple, Sally J Singh, David C Thomas, Mark Toshner, Aziz Sheikh, Christopher E Brightling, Louise V Wain, Rachael A Evans, the PHOSP-COVID Collaborative Group

## Abstract

- PHOSP-COVID is a national UK multi-centre cohort study of patients who were hospitalised for COVID-19 and subsequently discharged.
- PHOSP-COVID was established to investigate the medium- and long-term sequelae of severe COVID-19 requiring hospitalisation, understand the underlying mechanisms of these sequelae, evaluate the medium- and long-term effects of COVID-19 treatments, and to serve as a platform to enable future studies, including clinical trials.
- Data collected covered a wide range of physical measures, biological samples, and Patient Reported Outcome Measures (PROMs).
- Participants could join the cohort either in Tier 1 only with remote data collection using hospital records, a PROMs app and postal saliva sample for DNA, or in Tier 2 where they were invited to attend two specific research visits for further data collection and biological research sampling. These research visits occurred at five (range 2-7) months and 12 (range 10-14) months post-discharge. Participants could also participate in specific nested studies (Tier 3) at selected sites.
- All participants were asked to consent to further follow-up for 25 years via linkage to their electronic healthcare records and to be re-contacted for further research.
- In total, 7935 participants were recruited from 83 UK sites: 5238 to Tier 1 and 2697 to Tier 2, between August 2020 and March 2022.
- Cohort data are held in a Trusted Research Environment and samples stored in a central biobank. Data and samples can be accessed upon request and subject to approvals.

## Introduction

### Why was the cohort set up?

To date, there have been over 750 million reported cases of COVID-19 globally since the pandemic began in early 2020 (1). In the UK, there have been over one million patients hospitalised and 180,000 deaths due to COVID-19 (2). At the time of conception of the PHOSP-COVID cohort in March 2020, the longer-term pulmonary and multisystem effects of COVID-19 and impact on health status were unknown (3). We identified a need to establish a cohort of hospitalised COVID-19 survivors to collect detailed information about the medium- and long–term effects of COVID-19 on physical and mental health, lifestyle, and occupation status.

Although the majority of individuals with COVID-19 were not hospitalised, we expected the consequences of COVID-19 might be most pronounced after severe illness. Furthermore, the pressures on health systems during the pandemic needed to be taken into consideration when establishing a new clinical cohort. Therefore, we designed the PHOSP-COVID study to align with clinical follow-up reviews of hospitalised patients, where possible.

PHOSP-COVID was designed to take a patient-centred, holistic approach to understand the medium- and long-term effects of COVID-19 recognising the need to consider physical and mental health, social support, and lifestyle. There were three main aims of PHOSP-COVID:

1. To determine the medium- and long-term health (and health economic) sequelae of COVID-19 in post-hospitalisation survivors; to define demographic, clinical and molecular biomarkers of susceptibility, including to severity of the acute illness and development, progression, and resolution of sequelae.
2. To understand the impact of in-patient and post-discharge, pharmacological and non-pharmacological interventions on long-term sequelae of COVID-19.
3. To build the foundation for in-depth studies of emergent conditions and worsening of premorbid disease to inform precision medicine in at-risk groups by directing new clinical trials and care for current and future patients with long COVID.

### Who is in the cohort?

Individuals who were discharged from hospital between 1 February 2020 and 31 March 2021 were invited to participate in the PHOSP-COVID study if they were: aged 18 years or above, admitted to an admissions unit or ward at a participating UK hospital with confirmed or clinically suspected COVID-19, and were able to provide informed consent either personally or via a consultee or an appropriate representative. Exclusion criteria included: admission due to a confirmed diagnosis of a different pathogen with no indication or likelihood of co-infection with COVID-19, attendance to emergency department only, declined to provide informed consent or life-limiting illness with life expectancy less than six months such as disseminated malignancy. Patients were invited to participate by research teams based at the hospital site from which they were discharged: 83 sites from England, Northern Ireland, Scotland, and Wales participated. Different methods were used to obtain consent including: face-to-face, telephone, postal and eConsent.

Participants could join the cohort as Tier 1 participants only with remote extraction of information using hospital records and a PROMs app, and with a postal saliva sample for DNA, or could join as Tier 2 participants where they were invited to attend two research visits in addition to their clinical follow up for further data collection and biological research sampling (**Figure 1**).

**Figure 1:**
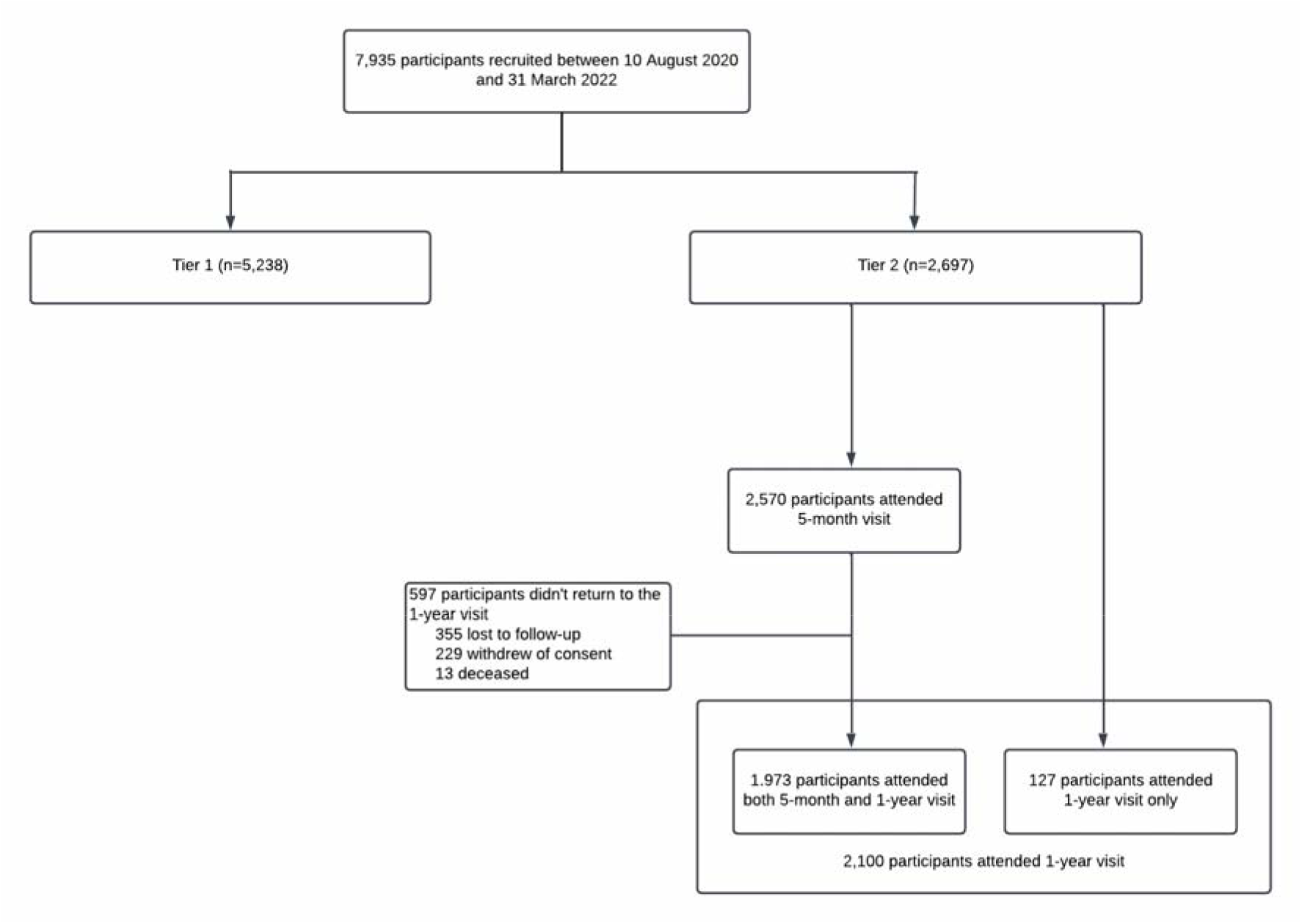
Consort diagram of the PHOSP-COVID study. * The wide range window for the first research visit (2-7 months) was deliberately chosen to accommodate the variation of planned clinical follow up appointments across the different participating sites and to allow the research visit to coincide with the planned clinical follow-up appointments.

Participants in either Tier 1 or Tier 2 could additionally join Tier 3 sub-studies where they were either recalled for additional research procedures or undertook additional research procedures during their Tier 2 research visits. For example, a subset of participants had an extended blood draw to enable additional sampling and advanced cellular studies (4) and another subset was invited to undertake up to three whole body magnetic resonance imaging (MRI) scans to examine the effect of COVID-19 on multiple body organs (Capturing MultiORgan Effects of COVID-19, C-MORE sub study) (5).

A total of 7935 participants were recruited into the PHOSP-COVID cohort, 5238 participants to Tier 1 and 2697 to Tier 2, between 10^th^ August 2020 and 31^st^ March 2022. The participants’ demographics, comorbidities, and admission characteristics are detailed in **Table 1** and **Table S1**. Of these, around 1000 participants to date have also been included in Tier 3 studies.

**Table 1:**
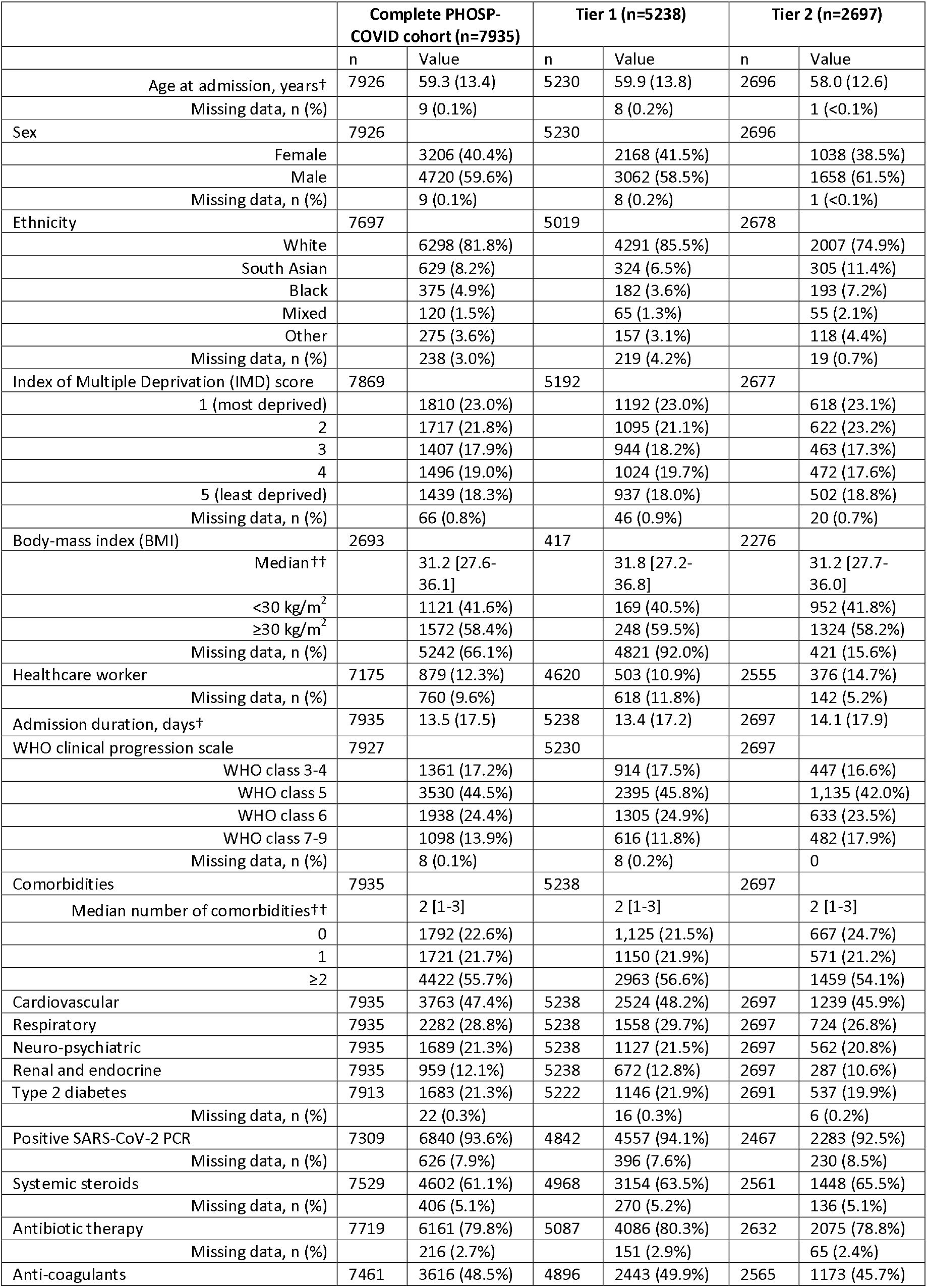
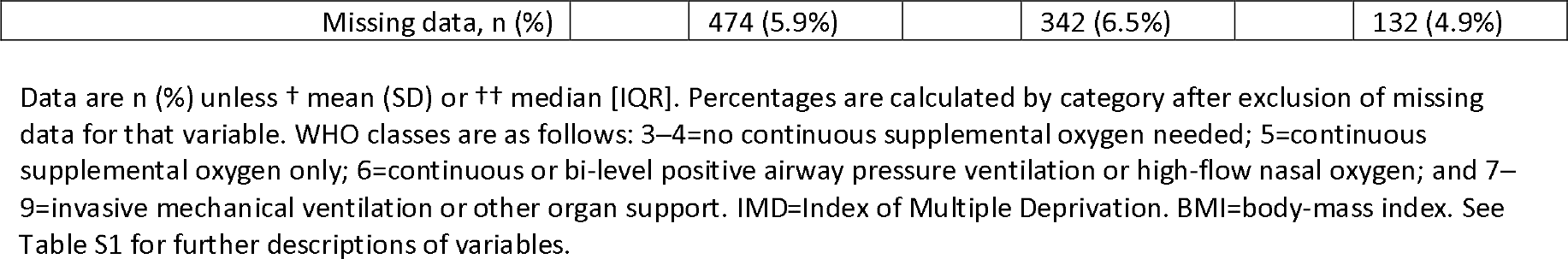
Participants demographics, comorbidities, and admission characteristics of the PHOSP-COVID cohort.

|  | Complete PHOSP-COVID cohort (n=7935) |  | Tier 1 (n=5238) |  | Tier 2 (n=2697) |  |
| --- | --- | --- | --- | --- | --- | --- |
|  | n | Value | n | Value | n | Value |
| Age at admission, yearst | 7926 | 59.3 (13.4) | 5230 | 59.9 (13.8) | 2696 | 58.0 (12.6) |
| Missing data, n (%) |  | 9 (0.1%) |  | 8 (0.2%) |  | 1 (<0.1%) |
| Sex | 7926 |  | 5230 |  | 2696 |  |
| Female |  | 3206 (40.4%) |  | 2168 (41.5%) |  | 1038 (38.5%) |
| Male |  | 4720 (59.6%) |  | 3062 (58.5%) |  | 1658 (61.5%) |
| Missing data, n (%) |  | 9 (0.1%) |  | 8 (0.2%) |  | 1 (<0.1%) |
| Ethnicity | 7697 |  | 5019 |  | 2678 |  |
| White |  | 6298 (81.8%) |  | 4291 (85.5%) |  | 2007 (74.9%) |
| South Asian |  | 629 (8.2%) |  | 324 (6.5%) |  | 305 (11.4%) |
| Black |  | 375 (4.9%) |  | 182 (3.6%) |  | 193 (7.2%) |
| Mixed |  | 120 (1.5%) |  | 65 (1.3%) |  | 55 (2.1%) |
| Other |  | 275 (3.6%) |  | 157 (3.1%) |  | 118 (4.4%) |
| Missing data, n (%) |  | 238 (3.0%) |  | 219 (4.2%) |  | 19 (0.7%) |
| Index of Multiple Deprivation (IMD) score | 7869 |  | 5192 |  | 2677 |  |
| 1 (most deprived) |  | 1810 (23.0%) |  | 1192 (23.0%) |  | 618 (23.1%) |
| 2 |  | 1717 (21.8%) |  | 1095 (21.1%) |  | 622 (23.2%) |
| 3 |  | 1407 (17.9%) |  | 944 (18.2%) |  | 463 (17.3%) |
| 4 |  | 1496 (19.0%) |  | 1024 (19.7%) |  | 472 (17.6%) |
| 5 (least deprived) |  | 1439 (18.3%) |  | 937 (18.0%) |  | 502 (18.8%) |
| Missing data, n (%) |  | 66 (0.8%) |  | 46 (0.9%) |  | 20 (0.7%) |
| Body-mass index (BMI) | 2693 |  | 417 |  | 2276 |  |
| Median†† |  | 31.2 [27.6-36.1] |  | 31.8 [27.2-36.8] |  | 31.2 [27.7-36.0] |
| <30 kg/m<sup>2</sup> |  | 1121 (41.6%) |  | 169 (40.5%) |  | 952 (41.8%) |
| ≥30 kg/m<sup>2</sup> |  | 1572 (58.4%) |  | 248 (59.5%) |  | 1324 (58.2%) |
| Missing data, n (%) |  | 5242 (66.1%) |  | 4821 (92.0%) |  | 421 (15.6%) |
| Healthcare worker | 7175 | 879 (12.3%) | 4620 | 503 (10.9%) | 2555 | 376 (14.7%) |
| Missing data, n (%) |  | 760 (9.6%) |  | 618 (11.8%) |  | 142 (5.2%) |
| Admission duration, dayst | 7935 | 13.5 (17.5) | 5238 | 13.4 (17.2) | 2697 | 14.1 (17.9) |
| WHO clinical progression scale | 7927 |  | 5230 |  | 2697 |  |
| WHO class 3-4 |  | 1361 (17.2%) |  | 914 (17.5%) |  | 447 (16.6%) |
| WHO class 5 |  | 3530 (44.5%) |  | 2395 (45.8%) |  | 1,135 (42.0%) |
| WHO class 6 |  | 1938 (24.4%) |  | 1305 (24.9%) |  | 633 (23.5%) |
| WHO class 7-9 |  | 1098 (13.9%) |  | 616 (11.8%) |  | 482 (17.9%) |
| Missing data, n (%) |  | 8 (0.1%) |  | 8 (0.2%) |  | 0 |
| Comorbidities | 7935 |  | 5238 |  | 2697 |  |
| Median number of comorbidities†† |  | 2 [1-3] |  | 2 [1-3] |  | 2 [1-3] |
| 0 |  | 1792 (22.6%) |  | 1,125 (21.5%) |  | 667 (24.7%) |
| 1 |  | 1721 (21.7%) |  | 1150 (21.9%) |  | 571 (21.2%) |
| ≥2 |  | 4422 (55.7%) |  | 2963 (56.6%) |  | 1459 (54.1%) |
| Cardiovascular | 7935 | 3763 (47.4%) | 5238 | 2524 (48.2%) | 2697 | 1239 (45.9%) |
| Respiratory | 7935 | 2282 (28.8%) | 5238 | 1558 (29.7%) | 2697 | 724 (26.8%) |
| Neuro-psychiatric | 7935 | 1689 (21.3%) | 5238 | 1127 (21.5%) | 2697 | 562 (20.8%) |
| Renal and endocrine | 7935 | 959 (12.1%) | 5238 | 672 (12.8%) | 2697 | 287 (10.6%) |
| Type 2 diabetes | 7913 | 1683 (21.3%) | 5222 | 1146 (21.9%) | 2691 | 537 (19.9%) |
| Missing data, n (%) |  | 22 (0.3%) |  | 16 (0.3%) |  | 6 (0.2%) |
| Positive SARS-CoV-2 PCR | 7309 | 6840 (93.6%) | 4842 | 4557 (94.1%) | 2467 | 2283 (92.5%) |
| Missing data, n (%) |  | 626 (7.9%) |  | 396 (7.6%) |  | 230 (8.5%) |
| Systemic steroids | 7529 | 4602 (61.1%) | 4968 | 3154 (63.5%) | 2561 | 1448 (65.5%) |
| Missing data, n (%) |  | 406 (5.1%) |  | 270 (5.2%) |  | 136 (5.1%) |
| Antibiotic therapy | 7719 | 6161 (79.8%) | 5087 | 4086 (80.3%) | 2632 | 2075 (78.8%) |
| Missing data, n (%) |  | 216 (2.7%) |  | 151 (2.9%) |  | 65 (2.4%) |
| Anti-coagulants | 7461 | 3616 (48.5%) | 4896 | 2443 (49.9%) | 2565 | 1173 (45.7%) |

Overall, the cohort has a mean age of 60 years, 40% of participants are female, 82% report white ethnicity and 23% are from the lowest quintile of the Index of Multiple Deprivation (IMD). The cohort was co-morbid with more than 55% of participants having two or more pre-existing comorbidities at the time of hospital admission. More than 90% had a positive SARS-CoV-2 RT-PCR test result on admission and 38% required non-invasive or invasive ventilation (Class 6 or above on the WHO clinical progression scale) (6) during their original hospital admission for COVID-19.

Given the pressures of the ongoing pandemic during recruitment, non-response to invitations to join the study was not recorded.

### How often have they been followed up?

Data collection for Tier 1 participants was restricted to available clinical data from routine hospital follow-up plus the collection of PROMs via an app every three months for up to one year post discharge. Tier 2 participants were invited to two research visits: the first between 2-7 months, and the second between 10-14 months post hospital discharge. Of the 2570 Tier 2 participants who attended the first research visit (labelled here as five-month visit due to median length of time between discharge and the visit), 1973 participants also attended a second research visit (labelled one-year visit). A further 127 Tier 2 participants attended the one-year visit only (**Figure 1**). The characteristics of the 597 participants who did not return for a one-year visit are listed in **Table S2**.

All participants provided consent for further data collection via linkage to retrospective and prospective health and social care records including primary care, Hospital Episode Statistics, pathology records, prescribing records, and specialist tertiary clinical databases for up to 25 years. Participants were also invited to provide consent to be re-contacted for further research, including Tier 3 sub-studies, such as mechanistic studies and clinical trials (7).

### What has been measured?

A summary of the data collected for PHOSP-COVID participants is provided in **Table 2**. For all participants, information about their demographics, acute illness and hospital admission were obtained retrospectively from the hospital notes. This included: comorbidities, presenting symptoms, length of stay, severity of acute illness assessed by the WHO clinical progression scale, treatment received, complications and common clinical test results. Hospital records were also reviewed to collect clinical data obtained from any planned follow-up appointments organised by the local hospital team after discharge. These included: physiological tests and imaging (e.g., electrocardiography, chest radiograph findings, etc.), routine blood test results and if available PROMs via clinical questionnaires assessing for example: health-related quality of life (HRQoL), clinical symptoms, and mental health (**Table SM1**). Further data were collected on post-discharge care accessed including mental health interventions, recovery/rehabilitation programmes and details from any emergency hospital admission for up to one year post discharge.

**Table 2:**
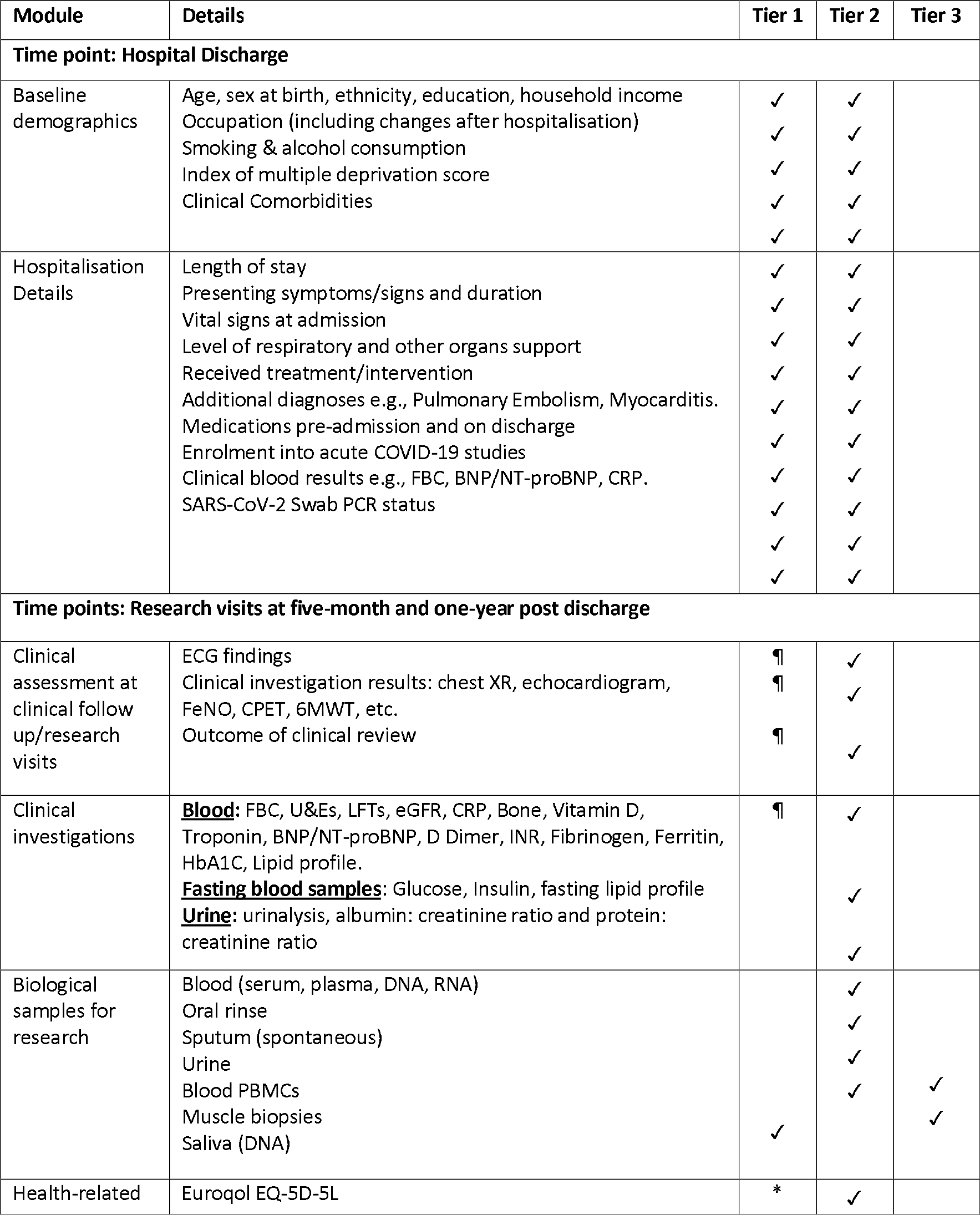
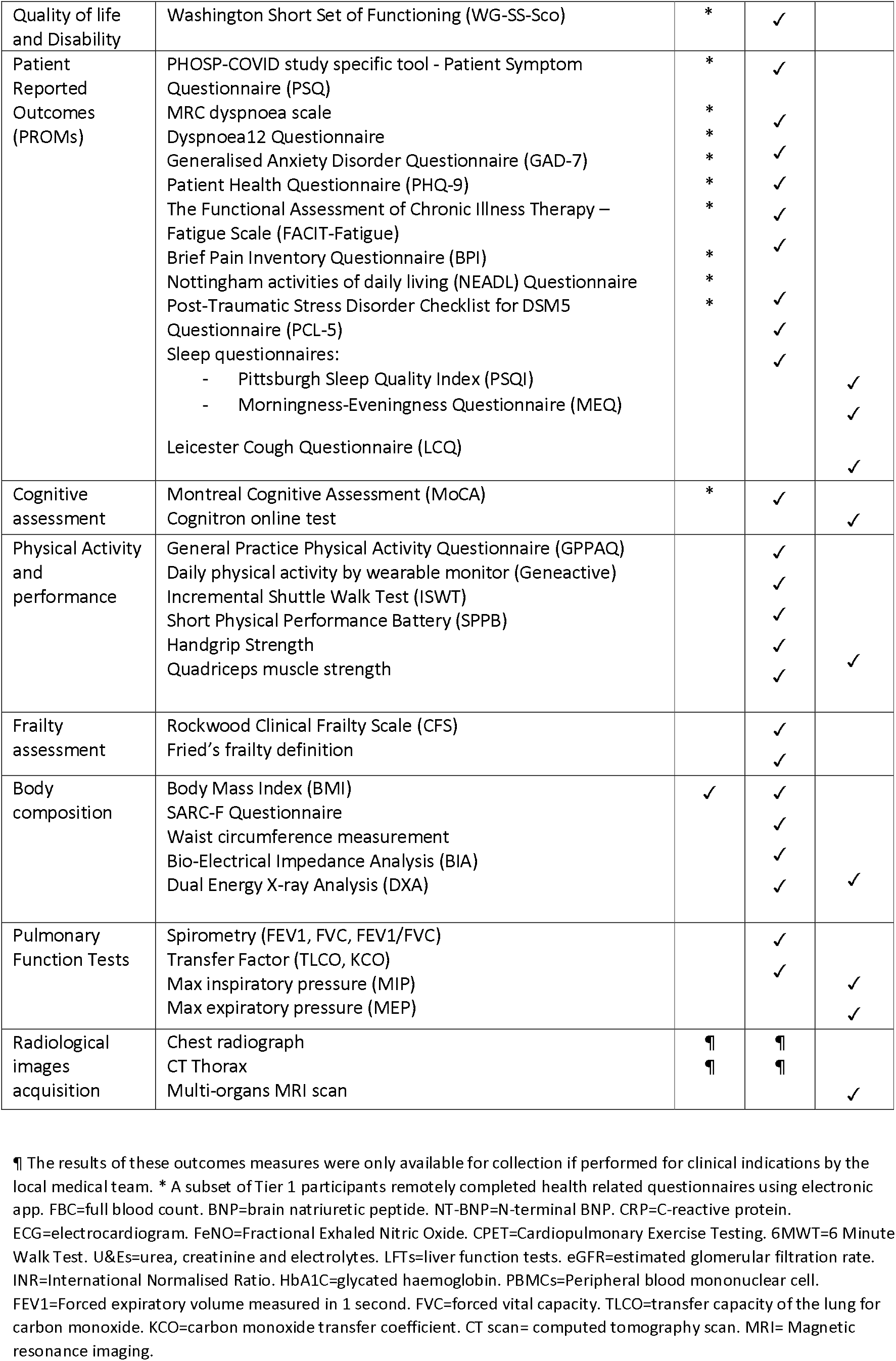
PHOSP-COVID outcome measures.

For participants in Tier 1, clinical data were obtained from medical records and no specific research visit was undertaken. However, a subset of Tier 1 participants used an online app to remotely complete PROM questionnaires and a bespoke study-specific Patient Symptom Questionnaire (PSQ) (**Appendix 1**). The PSQ was used to collect information about ongoing symptoms, changes in occupation, and perceived recovery where the participant was asked to answer ‘yes’, ‘no’ or ‘not sure’ to the question “Do you feel fully recovered from COVID-19?” (**Appendix 1**). A total of 371 participants provided 519 entries using the online PROMs app (142 Tier 1 and 229 Tier 2) between April 2021 and April 2022. Another subset of Tier 1 participants provided a saliva sample for DNA analysis via a collection kit posted to their home (**Table S3**).

At Tier 2 research visits, clinical questionnaires, procedures and sampling were undertaken including completion of the PSQ. Physical performance was assessed using questionnaires and physical tests including: handgrip and quadriceps strength, Short Physical Performance Battery (SPPB) and Incremental Shuttle Walk Test (ISWT). All Tier 2 participants were additionally invited to undertake daily physical activity monitoring using a wearable GENEactive© accelerometer for 14 days. Lung function was assessed using spirometry and measurement of gas transfer when feasible given the COVID-19 restrictions on aerosol generating procedures (**Table 3**).

**Table 3:**
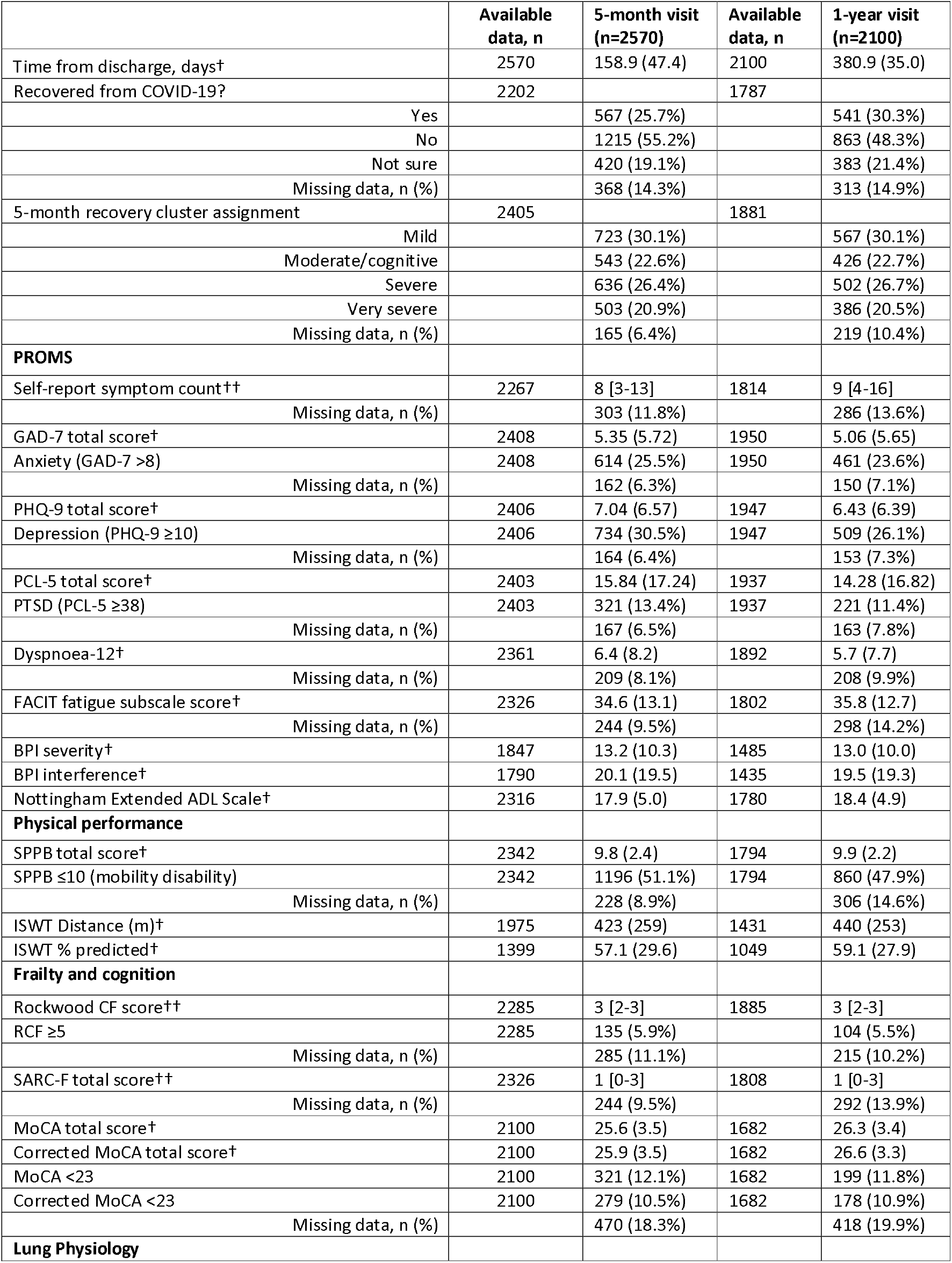
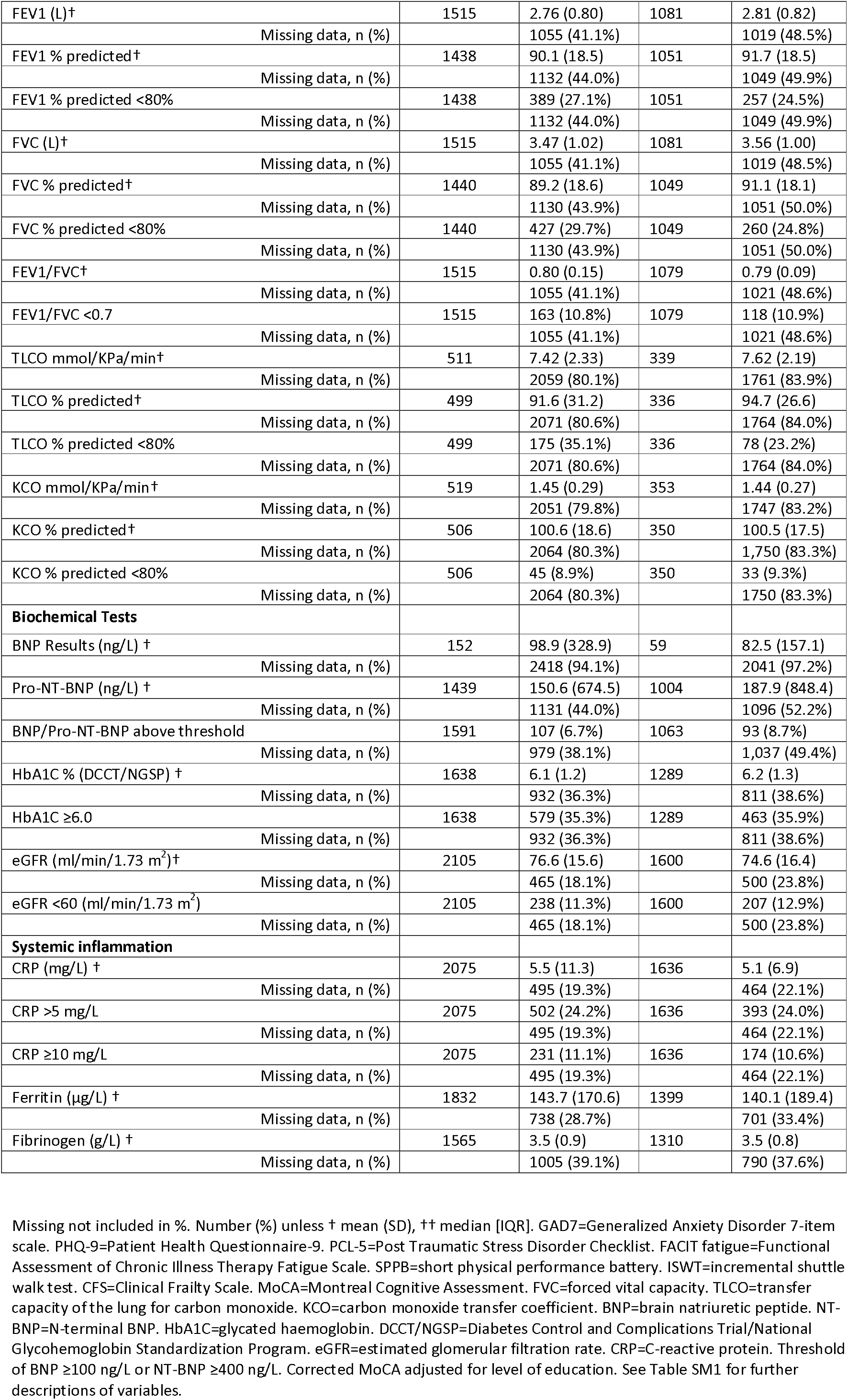
Patient-reported outcome measures, physiological and biochemical tests, among Tier 2 participants stratified by the research visits.

All assessments were performed as part of the two dedicated research visits except when relevant measures were already available from clinical follow-up appointments at the corresponding time points to reduce procedures burden and duplication.

All Tier 2 participants were invited to provide blood, urine, oral rinse, and sputum samples for research purposes. Six different blood sample tube types were used: plasma (EDTA, lithium heparin, citrate), serum, DNA and RNA (**Table S3**). All samples were minimally processed at the local site according to a core study laboratory standard operating procedure manual, before being shipped at intervals for longer-term storage at a central laboratory. This centralisation of samples facilitated their use in multi-site studies. Participants were asked to consent to use of their samples by other researchers, including commercial parties, both in the UK and abroad. Participants were given an option to decline their consent for genetic studies.

The participants’ consent to access healthcare records allowed access and acquisition of clinically indicated images including chest x-ray and thoracic CT scans from certain participating sites, which were transferred to a national imaging database (National COVID-19 Chest Imaging Database) for analysis and secure storage (**Table S4**).

Procedures for Tier 3 sub-studies were dependent on the specific criteria of the project e.g., whole body MRI imaging scans as part of C-MORE sub-study (**Table S5**), body composition measurements using Dual Energy X-ray Analysis (DXA) imaging or further cognitive assessment using the Cognitron online test (**Table 2**).

### What have we achieved?

#### Priority setting to identify 10 key research questions regarding the long-term sequelae of COVID-19

In order to ensure the patient voice was central to the research undertaken using the PHOSP-COVID cohort, a joint patient and clinician priority setting exercise was undertaken between December 2020 and March 2021 to determine 10 priority research questions (8). The priority setting incorporated views from adults with self-reported experience of long COVID (both hospitalised and non-hospitalised participants), carers, clinicians, and clinical researchers. A modified version of the James Lind Alliance (JLA) priority setting partnerships (PSP) process was used (9). A total of 119 initial questions were gathered prior to refining, rewording, and grouping into a shorter list of 24 questions which was shared through an online prioritisation survey receiving 882 responses (mainly from individuals with self-reported long COVID). The final top 10 research questions were agreed at a dedicated prioritisation workshop mediated by independent JLA facilitators and hosted via videoconference. The final top 10 research questions are listed in **Table S6**.

#### Key research findings to date

##### Significant burden of ongoing health impairment

Results from the first 1077 Tier 2 participants at five months post-discharge highlighted that only 29% of participants felt fully recovered, 20% reported a new disability assessed by the Washington Group Short Set on Functioning (WG-SS), and 18% were no longer working (10). The 10 most reported symptoms were: aching muscles, fatigue, physical slowing down, impaired sleep quality, joint pain or swelling, limb weakness, breathlessness, pain, short-term memory loss, and slowing down in thinking. These findings were consistent with reported symptoms from smaller cohorts or cohorts of patients with a less severe initial illness (11-13). Around one in four of the cohort had clinically relevant symptoms of anxiety and depression and nearly half of the participants had features of functional impairment measured using ISWT and SPPB at five months post-discharge. There was also evidence of specific organ impairment: 35% had prediabetes or diabetes, 31% had impaired lung function, 17% had at least mild cognitive impairment, 13% abnormal kidney function, and 7% raised brain natriuretic peptide (BNP). Further investigation of post-COVID residual lung abnormalities (RLA) using clinical thoracic imaging at a median of four months post discharge, revealed abnormalities affecting at least 10% of the lung were observed in 79.4% of a subset of 209 PHOSP-COVID participants (14). The prevalence of RLA was estimated between 8.5% and 11.7% and a proposed clinically applicable risk stratification suggested that 7.8% of the examined cohort had moderate- to very-high risk of RLA post-COVID hospitalisation.

A striking finding was the lack of a clear association between the severity of the acute illness and the ongoing symptoms, mental and physical health impairments with the exception of pulmonary function tests and walking performance, which were worse in the group who received invasive mechanical ventilation (10).

At one-year after hospital discharge there was very little improvement from five-months visit in self-perceived recovery, ongoing symptoms, mental health, physical performance, cognitive impairment, and organ impairment (15). The top 10 most prevalent symptoms were also similar to those at five-months. Frailty and pre-frailty were present in more than two-thirds of participants at one year (16). A fall in the number of participants working at one-year was seen with 8.5% of those who were working before hospitalisation no longer working and 34.6% of participants reporting that COVID-19 had resulted in a change in their occupation (**Table S7**). Results from the complete Tier 2 cohort for the early and one year research visits are included in **Tables 3 and 4**.

**Table 4:** Health-related quality of life and disability among Tier 2 participants stratified by the research visits.

|  | Available data, n | Pre-COVID (n=2697) | Available data, n | 5 months (n=2570) | Available data, n | 1 year (n=2100) |
| --- | --- | --- | --- | --- | --- | --- |
| EQ-5D-5L utility index† | 2170 | 0.82 (0.23) | 2113 | 0.71 (0.25) | 1740 | 0.71 (0.25) |
| Missing data, n (%) |  | 527 (19.5%) |  | 457 (17.8%) |  | 360 (17.1%) |
| EQ-5D-5L utility index delta change† | - | - | 1757 | -0.11 (0.22) | 1498 | -0.11 (0.22) |
| Missing data, n (%) |  |  |  | 813 (31.6%) |  | 602 (28.7%) |
| EQ-5D-5L VAS† | 2095 | 79.5 (17.5) | 2106 | 70.1 (20.0) | 1731 | 70.4 (20.6) |
| Missing data, n (%) |  | 602 (22.3%) |  | 464 (18.1%) |  | 369 (17.6%) |
| EQ-5D-5L VAS delta change† | - | - | 1697 | - 9.9 (19.4) | 1435 | -9.8 (19.8) |
| Missing data, n (%) |  |  |  | 873 (33.9%) |  | 665 (31.7%) |
| WG-SS-SCo | - | - | 2208 | 532 (24.1%) | 1793 | 389 (21.7%) |
| Missing data, n (%) |  |  |  | 362 (14.1%) |  | 307 (14.6%) |
| WG-SS-SCo new disability | - | - | 1659 | 317 (19.1%) | 491 | 93 (18.9%) |
| Missing data, n (%) |  |  |  | 911 (35.5%) |  | 1609 (76.6%) |
| PSQ Breathlessness†† | 2162 | 0 [0-2] | 2193 | 4 [1-6] | 1770 | 2 [0-5] |
| Missing data, n (%) |  | 535 (19.8%) |  | 377 (14.7%) |  | 330 (15.7%) |
| PSQ Cough†† | 2153 | 0 [0-1] | 2184 | 1 [0-4] | 1763 | 0 [0-2] |
| Missing data, n (%) |  | 544 (20.2%) |  | 386 (15.0%) |  | 337 (16.0%) |
| PSQ Fatigue†† | 2152 | 0 [0-2] | 2183 | 5 [2-7] | 1765 | 3 [1-6] |
| Missing data, n (%) |  | 545 (20.2%) |  | 387 (15.1%) |  | 335 (15.9%) |
| PSQ Poor Sleep†† | 2151 | 1 [0-4] | 2177 | 4 [1-7] | 1766 | 3 [0-6] |
| Missing data, n (%) |  | 546 (20.2%) |  | 393 (15.3%) |  | 334 (15.9%) |
| PSQ Pain†† | 2138 | 0 [0-3] | 2169 | 3 [0-6] | 1763 | 2 [0-5] |
| Missing data, n (%) |  | 559 (20.7%) |  | 401 (15.6%) |  | 337 (16.0%) |

Missing not included in %. Number (%) unless † mean (SD), †† median [IQR]. EQ-5D-5L VAS = Euroqol five level visual analogue scale 0-100. WG-SS-SCo = Washington Group Short Set of Functioning Severity Continuum. PSQ = Patient Symptoms Questionnaires. See Table SM1 for further descriptions of variables.

##### Risk factors for lack of recovery

The risk factors associated with lack of recovery at one-year were: being female, being obese and having received invasive mechanical ventilation during the acute illness (WHO clinical progression scale class 7-9) (15). History of treatment with acute corticosteroids during the acute admission was not associated with any effect on patient perceived recovery at one-year despite the beneficial acute effects (17). Frailty was also positively associated with non-recovery and reduced health-related quality of life at one year following discharge (16).

We identified risk factors for new or worse breathlessness post-COVID at five months including socio-economic deprivation, pre-existing depression/anxiety, female sex and longer hospital stay (18). Further analysis has also revealed disrupted sleep, present in 62% of the cohort, associated with dyspnoea, anxiety, and muscle weakness revealing an intriguing potential therapeutic intervention (19).

##### Recovery trajectory clusters

We undertook unsupervised cluster modelling using validated objective measures of breathlessness, fatigue, anxiety, depression, post-traumatic stress disorder (PTSD), physical performance and cognitive impairments at five months (10) and one year post-hospital discharge (15), and described four ‘recovery clusters’. The severity of most of the health impairments largely tracked together in the ‘very severe’, ‘severe’ and ‘mild’ clusters whilst the ‘moderate’ cluster was dominated by cognitive impairment (**Figure 2**). The more severe clusters were associated with female sex, higher body-mass index (BMI), a higher number of symptoms, reduced physical function and elevated C-reactive protein levels. The ‘very severe’ recovery cluster was associated with fewer days/week containing continuous bouts of moderate-to-vigorous physical activity, longer total sleep time, and higher variability in sleep timing (20). Although these are associations for which causal directions of effect have not been determined, these data highlight potential therapeutic targets (21).

**Figure 2:**
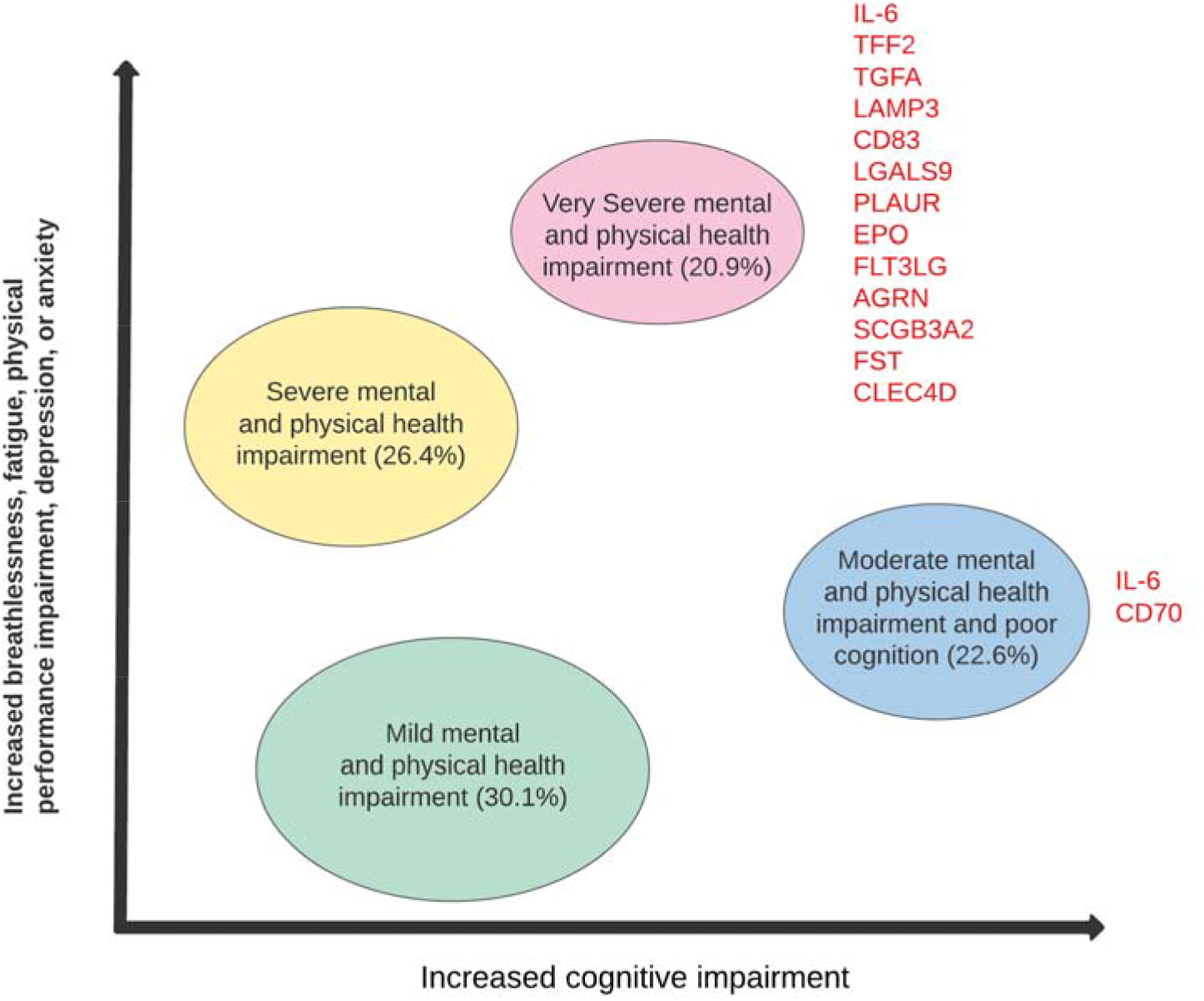
Illustration of the four cluster phenotypes mental, cognitive, and physical health impairments with associated inflammatory biomarkers. The figure shows the distribution of the four recovery cluster phenotypes and the list of identified proteins that were significantly differentially expressed (compared with the reference mild cluster) after FDR adjustment. FDR=false detection rate. IL-6=interleukin-6. TFF2=trefoil factor 2. TGFA=transforming growth factor α. LAMP3=lysosomal associated membrane protein 3. CD83=CD83 molecule. LGALS9=galectin-9. PLAUR=urokinase plasminogen activator surface receptor. EPO=erythropoietin. FLT3LG=FMS-related receptor tyrosine kinase 3 ligand. AGRN=agrin. SCGB3A2=secretoglobin family 3A member 2. FST=follistatin. CLEC4D=C-type lectin domain family 4 member D. CD70=CD70 molecule. Adapted from “Physical, cognitive, and mental health impacts of COVID-19 after hospitalisation (PHOSP-COVID): a UK multicentre, prospective cohort study” by Evans RA, et al., 2021, *The Lancet Respiratory Medicine*. 2021;9(11):1275-87. Licensed under the Creative Commons Attribution (CC BY 4.0).

To investigate the inflammatory response further, levels of 296 inflammatory proteins were measured at five months post-discharge. Thirteen proteins, including IL-6, were elevated in the ‘very severe’ and the ‘moderate with cognitive impairment’ clusters compared with the ‘mild cluster’ (**Figure 2**). These mediators of tissue damage and repair provide plausible biological mechanisms behind the symptoms and health impairments associated with severe long COVID (15).

#### Strengths and limitations

The large number of clinical variables collected, coupled with the biological research sampling, makes PHOSP-COVID one of the largest deeply-phenotyped cohorts of hospitalised COVID-19 survivors in the world. Cross-sectional and longitudinal multi-omics markers are being measured in Tier 2 participants (for example, plasma inflammatory proteomics data are already available for around 1000 participants). These may uncover underlying mechanistic pathways implicated in long COVID pathology and inform interventional trials. We have linked participants in PHOSP-COVID to the ISARIC study data, where applicable (22). This provides additional information relating to their hospitalisation as well as enabling linkage to samples taken during acute hospital admission. This linkage is also being developed to facilitate linkage to other resources including vaccine data, viral strain data and electronic healthcare records e.g., OpenSAFELY (in progress at the time of writing).

The multi-dimensional results generated by the PHOSP-COVID cohort are helping to shape and prioritise provision of clinical care at times where the national health services both locally and globally are under significant pressure after the pandemic (23). Setting priority research questions and identifying risk groups will focus the efforts of both clinical and academic institutions at managing the large volume of patients with long COVID (8, 24).

The study was designed as a cohort with the study population being defined by hospitalisation due to COVID-19 (and subsequent discharge alive from hospital) and a range of outcomes captured enabling nested case-control analyses. As such, no external comparator groups (i.e., non-hospitalised COVID-19 survivors, individuals hospitalised with other viral infections) were recruited to the study. However, this has been partially mitigated by using external cohorts or healthy controls to examine certain hypotheses (19, 25).

As participants were prospectively recruited following discharge from hospital, data pertaining to pre-COVID-19 health status were only available from healthcare records or by participant recall introducing the potential for recall bias. There is also unavoidable selection bias as some of the participants might have accepted the invitation to the study due to the severity of their ongoing symptoms. This is particularly relevant to Tier 2 participants who were younger, more ethnically diverse, less comorbid and required more respiratory support compared to the participants included in the ISARIC4C consortium outputs, which are likely more representative of the overall hospitalised population in the UK (26, 27). However, the linkage to ISARIC and other public databases may help quantify and partially mitigate this bias.

As the PHOSP-COVID cohort included participants from 83 different sites and due to the pressure associated with providing clinical and academic services during the heights of the pandemic, there were considerable variations in the availability of collected data across these multiple sites. However, the large number of recruited participants still makes the PHOSP-COVID one of the largest multi-centre cohorts globally.

As recruitment began in August 2020, the cohort represents mainly patients who were admitted to hospital during the first year of the pandemic and so mostly preceded the emergence of the delta and omicron SARS-CoV2 variants and current guidance for acute therapeutics. In addition, as vaccination in the UK did not begin until late 2020, a large proportion of the cohort were vaccine naïve at initial hospital admission and at the five-month follow-up.

#### Where can I find out more and how can I access the data?

The PHOSP-COVID study website (https://www.phosp.org) contains an overview of the study, resources, information about people involved, and publications. Research activity using the study is organised across a series of Working Groups (**Figure 3**). These were established at the outset of the study to coordinate research, minimise duplication of efforts, and facilitate communication across research and clinical specialties. Researchers interested in undertaking research using PHOSP-COVID are encouraged to contact the relevant Working Group leads (https://www.phosp.org/working-group/) in the first instance. The data are currently held in the Outbreak Data Analysis Platform (ODAP, https://odap.ac.uk/). Researchers seeking to access these data are directed to https://www.phosp.org/resource/ for information and forms. Correspondence to be directed to Dr Rachael A Evans, the Co-Principal Investigator of PHOSP-COVID study.

**Figure 3:**
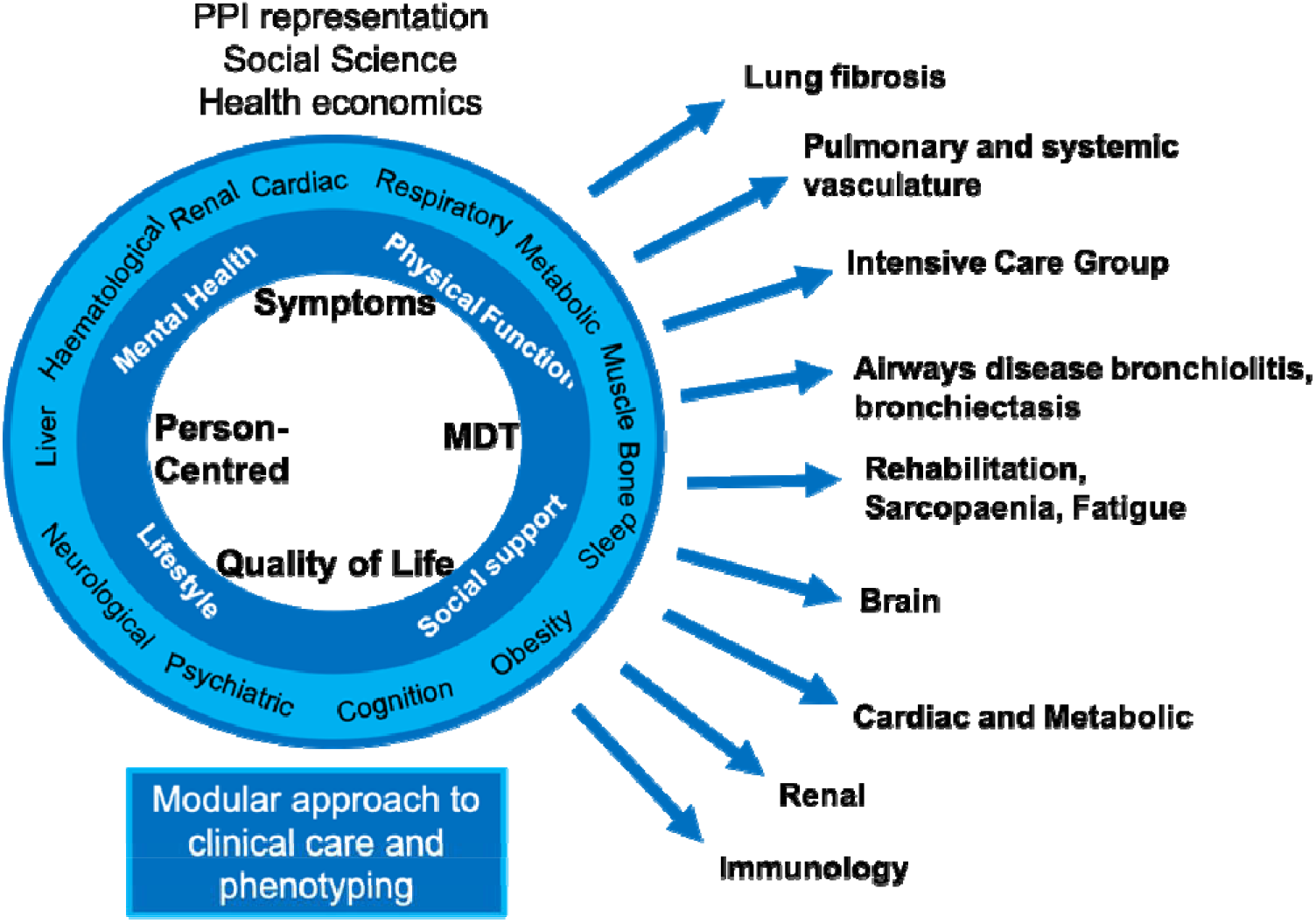
Modular approach to the clinical care and phenotyping with the PHOSP-COVID consortium different working groups. MDT=Multidisciplinary team. PPI=Patient and public involvement.

## Data Availability

The PHOSP-COVID study website (https://www.phosp.org) contains an overview of the study, resources, information about people involved, and publications. Research activity using the study is organised across a series of Working Groups (Figure 3). These were established at the outset of the study to coordinate research, minimise duplication of efforts, and facilitate communication across research and clinical specialties. Researchers interested in undertaking research using PHOSP-COVID are encouraged to contact the relevant Working Group leads (https://www.phosp.org/working-group/) in the first instance. The data are currently held in the Outbreak Data Analysis Platform (ODAP, https://odap.ac.uk/). Researchers seeking to access these data are directed to https://www.phosp.org/resource/ for information and forms. Correspondence to be directed to Dr Rachael A Evans, the Co-Principal Investigator of PHOSP-COVID study.

## Ethics approval

The study was approved by the Leeds West Research Ethics Committee (20/YH/0225) and is registered on the ISRCTN Registry (ISRCTN10980107).

## Author contributions

The manuscript was initially drafted by OE, RAE, and LVW, and further developed by the writing committee. CEB, RAE, LVW, JDC, L-PH, AH, MM, KP, BR, OE, HJCM, OCL, MR, AShi, ASi, MS, RMS, NJG, VCH, LH-W and AShe made substantial contributions to the conception and design of the work. LGH, KEL, RA, PB, CEBo, JSB, GC, NDB, NE, CE, JF, NH, JRH, MGJ, DP, PP, NMR, SLR-J, AART, CJ AMS and DGW made substantial contributions to the acquisition of data. All authors contributed to data interpretation, critical review, and revision of the manuscript. OE, HJCM, OCL have accessed and verified the underlying data. OE, RAE, CEB, and LVW were responsible for the decision to submit the manuscript, and are accountable for all aspects of the work in ensuring that questions related to the accuracy or integrity of any part of the work are appropriately investigated and resolved.

## Supplementary data

Supplementary data are available at IJE online.

## Acknowledgements

This study would not be possible without all the participants who have given their time and support. We thank all the participants and their families. We thank the many research administrators, health-care and social-care professionals who contributed to setting up and delivering the study at all of the 65 NHS trusts/Health boards and 25 research institutions across the UK, as well as all the supporting staff at the NIHR Clinical Research Network, Health Research Authority, Research Ethics Committee, Department of Health and Social Care, Public Health Scotland, and Public Health England, and support from the ISARIC Coronavirus Clinical Characterisation Consortium. We thank Kate Holmes at the NIHR Office for Clinical Research Infrastructure (NOCRI) for her support in coordinating the charities group. The PHOSP-COVID industry framework was formed to provide advice and support in commercial discussions, and we thank the Association of the British Pharmaceutical Industry as well NOCRI for coordinating this. We are very grateful to all the charities that have provided insight to the study: Action Pulmonary Fibrosis, Alzheimer’s Research UK, Asthma + Lung UK, British Heart Foundation, Diabetes UK, Cystic Fibrosis Trust, Kidney Research UK, MQ Mental Health, Muscular Dystrophy UK, Stroke Association Blood Cancer UK, McPin Foundations, Versus Arthritis and The Wolfsen Foundation. We thank the NIHR Leicester Biomedical Research Centre patient and public involvement group and Long Covid Support. This research was funded in whole or in part by the Wellcome Trust [209553/Z/17/Z]. For the purpose of open access, the author has applied a CC-BY public copyright licence to any author accepted manuscript version arising from this submission.

## Notes

**The PHOSP-COVID Collaborative Group:**

**Core Management Group**

*Chief Investigator* C E Brightling, *Members:* R A Evans (Lead Co-I), L V Wain (Lead Co-I), J D Chalmers, V C Harris, L P Ho, A Horsley, M Marks, K Poinasamy, B Raman, A Shikotra, A Singapuri.

**PHOSP-COVID Study Central Coordinating Team**

C E Brightling (Chief Investigator), R A Evans (*Lead Co-I*), L V Wain (*Lead Co-I*), R Dowling, C Edwardson, O Elneima, S Finney, N J Greening, B Hargadon, V Harris, L Houchen--Wolloff, O C Leavy, H J C McAuley, C Overton, T Plekhanova, R M Saunders, M Sereno, A Singapuri, A Shikotra, C Taylor, S Terry, C Tong, B Zhao.

**Steering Committee**

*Co-chairs:* D Lomas, E Sapey*, Institution representatives:* C Berry, C E Bolton, N Brunskill, E R Chilvers, R Djukanovic, Y Ellis, D Forton, N French, J George, N A Hanley, N Hart, L McGarvey, N Maskell, H McShane, M Parkes, D Peckham, P Pfeffer, A Sayer, A Sheikh, A A R Thompson, N Williams and core management group representation.

**Platforms**

**Bioresource**

W Greenhalf (*Co-Lead*), M G Semple (*Co-Lead*), M Ashworth, H E Hardwick, L Lavelle-Langham, W Reynolds, M Sereno, R M Saunders, A Singapuri, V Shaw, A Shikotra, B Venson, L V Wain.

**Data Hub**

A B Docherty (*Co-Lead*), E M Harrison (*Co-Lead*), A Sheikh (*Co-Lead*), J K Baillie, C E Brightling, L Daines, R Free, R A Evans, S Kerr, O C Leavy, N I Lone, H J C McAuley, R Pius, J K Quint, M Richardson, M Sereno, M Thorpe, L V Wain.

**Imaging Alliance**

M Halling-Brown (*Co-Lead*), F Gleeson (*Co-Lead*), J Jacob (*Co-Lead*), S Neubauer (*Co-Lead*) B Raman (*Co-Lead*) S Siddiqui (*Co-Lead*) J M Wild (*Co-Lead*), S Aslani, P Jezzard, H Lamlum, W Lilaonitkul, E Tunnicliffe, J Willoughby.

**Omics**

L V Wain (*Co-Lead)*, J K Baillie (*Co-Lead*), H Baxendale, C E Brightling, M Brown, J D Chalmers, R A Evans, B Gooptu, W Greenhalf, H E Hardwick, R G Jenkins, D Jones, I Koychev, C Langenberg, A Lawrie, P L Molyneaux, A Shikotra, J Pearl, M Ralser, N Sattar, R M Saunders, J T Scott, T Shaw, D Thomas, D Wilkinson.

**Working Groups**

**Airways**

L G Heaney (*Co-Lead*), A De Soyza (*Co-Lead*), C E Brightling, J S Brown, J Busby, J D Chalmers, C Echevarria, L Daines, O Elneima, R A Evans, J R Hurst, P Novotny, P Pfeffer, K Poinasamy, J K Quint, M Shankar-Hari, A Sheikh, S Siddiqui, S Walker, B Zheng.

**Brain**

J R Geddes (*Lead*), M Hotopf *(Co-Lead),* K Abel, R Ahmed, L Allan, C Armour, D Baguley, D Baldwin, C Ballard, K Bhui, G Breen, M Broome, T Brugha, E Bullmore, D Burn, F Callard, J Cavanagh, T Chalder, D Clark, A David, B Deakin, H Dobson, B Elliott, J Evans, R Francis, E Guthrie, P Harrison, M Henderson, A Hosseini, N Huneke, M Husain, T Jackson, I Jones, T Kabir, P Kitterick, A Korszun, I Koychev, J Kwan, A Lingford-Hughes, P Mansoori, H McAllister-Williams, K McIvor, L Milligan, R Morriss, E Mukaetova-Ladinska, K Munro, A Nevado-Holgado, T Nicholson, S Paddick, C Pariante, J Pimm, K Saunders, M Sharpe, G Simons, R Upthegrove, S Wessely.

**Cardiac**

G P McCann (*Lead*), S Amoils, C Antoniades, A Banerjee, R Bell, A Bularga, C Berry, P Chowienczyk, J P Greenwood, A D Hughes, K Khunti, L Kingham, C Lawson, K Mangion, N L Mills, A J Moss, S Neubauer, B Raman, A N Sattar, C L Sudlow, M Toshner.

**Immunology**

P J M Openshaw (*Lead*), D Altmann, J K Baillie, R Batterham, H Baxendale, N Bishop, C E Brightling, P C Calder, R A Evans, J L Heeney, T Hussell, P Klenerman, J M Lord, P Moss, S L Rowland-Jones, W Schwaeble, M G Semple, R S Thwaites, L Turtle, L V Wain, S Walmsley, D Wraith.

**Intensive Care**

M J Rowland (*Lead*), A Rostron (*Co-Lead*), J K Baillie, B Connolly, A B Docherty, N I Lone, D F McAuley, D Parekh, A Rostron, J Simpson, C Summers.

**Lung Fibrosis**

R G Jenkins (*Co-Lead*), J Porter (*Co-Lead*), R J Allen, R Aul, J K Baillie, S Barratt, P Beirne, J Blaikley, R C Chambers, N Chaudhuri, C Coleman, E Denneny, L Fabbri, P M George, M Gibbons, F Gleeson, B Gooptu, B Guillen Guio, I Hall, N A Hanley, L P Ho, E Hufton, J Jacob, I Jarrold, G Jenkins, S Johnson, M G Jones, S Jones, F Khan, P Mehta, J Mitchell, P L Molyneaux, J E Pearl, K Piper Hanley, K Poinasamy, J K Quint, D Parekh, P Rivera-Ortega, L C Saunders, M G Semple, J Simpson, D Smith, M Spears, L G Spencer, S Stanel, I Stewart, A A R Thompson, D Thickett, R Thwaites, L V Wain, S Walker, S Walsh, J M Wild, D G Wootton, L Wright.

**Metabolic**

S Heller (*Co-Lead*), M J Davies (*Co-Lead*), H Atkins, S Bain, J Dennis, K Ismail, D Johnston, P Kar, K Khunti, C Langenberg, P McArdle, A McGovern, T Peto, J Petrie, E Robertson, N Sattar, K Shah, J Valabhji, B Young.

**Pulmonary and Systematic Vasculature**

L S Howard (*Co-Lead*), Mark Toshner (*Co-Lead*), C Berry, P Chowienczyk, D Lasserson, A Lawrie, J Mitchell, L Price, J Rossdale, N Sattar, C Sudlow, A A R Thompson, J M Wild, M Wilkins.

**Rehabilitation, Sarcopenia and Fatigue**

S J Singh (*Co-Lead*), W D-C Man (*Co-Lead*), J M Lord (*Co-Lead*), N J Greening (*Co-Lead*), T Chalder (*Co-Lead*), J T Scott (*Co-Lead*), N Armstrong, E Baldry, M Baldwin, N Basu, M Beadsworth, L Bishop, C E Bolton, A Briggs, M Buch, G Carson, J Cavanagh, H Chinoy, E Daynes, S Defres, R A Evans, P Greenhaff, S Greenwood, M Harvie, M Husain, S MacDonald, A McArdle, H J C McAuley, A McMahon, M McNarry, C Nolan, K O’Donnell, D Parekh, Pimm, J Sargent, L Sigfrid, M Steiner, D Stensel, A L Tan, J Whitney, D Wilkinson, D Wilson, M Witham, D G Wootton, T Yates.

**Renal**

D Thomas (*Lead*), N Brunskill (*Co-Lead*), S Francis (*Co-Lead*), S Greenwood (*Co-Lead*), C Laing (*Co-Lead*), K Bramham, P Chowdhury, A Frankel, L Lightstone, S McAdoo, K McCafferty, M Ostermann, N Selby, C Sharpe, M Willicombe.

**Local Clinical Centre PHOSP-COVID trial staff** (Listed in alphabetical order)

**Airedale NHS Foundation Trust**

A Shaw (Principal Investigator: PI), L Armstrong, B Hairsine, H Henson, C Kurasz, L Shenton.

**Aneurin Bevan University Health Board**

S Fairbairn (PI), A Dell, N Hawkings, J Haworth, M Hoare, A Lucey, V Lewis, G Mallison, H Nassa, C Pennington, A Price, C Price, A Storrie, G Willis, S Young.

**Barts Health NHS Trust & Queen Mary University of London**

P Pfeffer (PI), K Chong-James, C David, W Y James, A Martineau, O Zongo.

**Barnsley Hospital NHS Foundation Trust**

A Sanderson (PI).

**Belfast Health and Social Care Trust & Queen’s University Belfast**

L G Heaney (PI), C Armour, V Brown, T Craig, S Drain, B King, N Magee, D McAulay, E Major, L McGarvey, J McGinness, R Stone.

**Betsi Cadwaladr University Health Board**

A Haggar (PI), A Bolger, F Davies, J Lewis, A Lloyd, R Manley, E McIvor, D Menzies, K Roberts, W Saxon, D Southern, C Subbe, V Whitehead.

**Borders General Hospital, NHS Borders**

H El-Taweel (PI), J Dawson, L Robinson.

**Bradford Teaching Hospitals NHS Foundation Trust**

D Saralaya (PI), L Brear, K Regan, K Storton.

**Cambridge University Hospitals NHS Foundation Trust, NIHR Cambridge Clinical Research Facility & University of Cambridge**

J Fuld (PI), A Bermperi, I Cruz, K Dempsey, A Elmer, H Jones, S Jose, S Marciniak, M Parkes, C Ribeiro, J Taylor, M Toshner, L Watson, J Worsley.

**Cardiff and Vale University Health Board**

R Sabit (PI), L Broad, A Buttress, T Evans, M Haynes, L Jones, L Knibbs, A McQueen, C Oliver, K Paradowski, J Williams.

**Chesterfield Royal Hospital NHS Trust**

E Harris (PI), C Sampson.

**Cwm Taf Morgannwg University Health Board**

C Lynch (PI), E Davies, C Evenden, A Hancock, K Hancock, M Rees, L Roche, N Stroud, T Thomas-Woods.

**East Cheshire NHS Trust**

M Babores (PI), J Bradley-Potts, M Holland, N Keenan, S Shashaa, H Wassall.

**East Kent Hospitals University NHS Foundation Trust**

E Beranova (PI), H Weston (PI), T Cosier, L Austin, J Deery, T Hazelton, C Price, H Ramos, R Solly, S Turney.

**Gateshead NHS Trust**

L Pearce (PI), W McCormack, S Pugmire, W Stoker, A Wilson.

**Guy’s and St Thomas’ NHS Foundation Trust**

N Hart (PI), LA Aguilar Jimenez, G Arbane, S Betts, K Bisnauthsing, A Dewar, P Chowdhury, A Dewar, G Kaltsakas, H Kerslake, MM Magtoto, P Marino, LM Martinez, M Ostermann, J Rossdale, TS Solano, E Wynn.

**Hampshire Hospitals NHS Foundation Trust**

N Williams (PI), W Storrar (PI), M Alvarez Corral, A Arias, E Bevan, D Griffin, J Martin, J Owen, S Payne, A Prabhu, A Reed, C Wrey Brown.

**Harrogate and District NHD Foundation Trust**

C Lawson (PI), T Burdett, J Featherstone, A Layton, C Mills, L Stephenson.

**Hull University Teaching Hospitals NHS Trust & University of Hull**

N Easom (PI), P Atkin, K Brindle, M G Crooks, K Drury, R Flockton, L Holdsworth, A Richards, D L Sykes, S Thackray-Nocera, C Wright.

**Hywel Dda University Health Board**

K E Lewis (PI), A Mohamed (PI), G Ross (PI), S Coetzee, K Davies, R Hughes, R Loosley, L O’Brien, Z Omar, H McGuinness, E Perkins, J Phipps, A Taylor, H Tench, R Wolf-Roberts.

**Imperial College Healthcare NHS Trust & Imperial College London**

L S Howard (PI), O Kon (PI), D C Thomas (PI), S Anifowose, L Burden, E Calvelo, B Card, C Carr, E R Chilvers, D Copeland, P Cullinan, P Daly, L Evison, T Fayzan, H Gordon, S Haq, R G Jenkins, C King, K March, M Mariveles, L McLeavey, N Mohamed, S Moriera, U Munawar, J Nunag, U Nwanguma, L Orriss-Dib, A Ross, M Roy, E Russell, K Samuel, J Schronce, N Simpson, L Tarusan, C Wood, N Yasmin.

**Kettering General Hospital NHS Trust**

R Reddy (PI), A-M Guerdette, M Hewitt, K Warwick, S White.

**King’s College Hospital NHS Foundation Trust & Kings College London**

A M Shah (PI), C J Jolley (PI), O Adeyemi, R Adrego, H Assefa-Kebede, J Breeze, M Brown, S Byrne, T Chalder, P Dulawan, N Hart, A Hayday, A Hoare, A Knighton, M Malim, S Patale, I Peralta, N Powell, A Ramos, K Shevket, F Speranza, A Te.

**Leeds Teaching Hospitals & University of Leeds**

P Beirne (PI), A Ashworth, J Clarke, C Coupland, M Dalton, E Wade, C Favager, J Greenwood, J Glossop, L Hall, T Hardy, A Humphries, J Murira, D Peckham, S Plein, J Rangeley, G Saalmink, A L Tan, B Whittam, N Window, J Woods.

**Lewisham & Greenwich NHS Trust**

G Coakley (PI).

**Liverpool University Hospitals NHS Foundation Trust & University of Liverpool**

D G Wootton (PI), L Turtle (PI), L Allerton, AM All, M Beadsworth, A Berridge, J Brown, S Cooper, A Cross, S Defres, S L Dobson, J Earley, N French, W Greenhalf, H E Hardwick, K Hainey, J Hawkes, V Highett, S Kaprowska, AL Key, L Lavelle-Langham, N Lewis-Burke, G Madzamba, F Malein, S Marsh, C Mears, L Melling, M J Noonan, L Poll, J Pratt, E Richardson, A Rowe, M G Semple, V Shaw, K A Tripp, L O Wajero, S A Williams-Howard, J Wyles.

**London North West University Healthcare NHS Trust**

S N Diwanji (PI), P Papineni (PI), S Gurram, S Quaid, G F Tiongson, E Watson.

**Manchester University NHS Foundation Trust & University of Manchester**

B Al-Sheklly (PI), A Horsley (PI), C Avram, J Blaikely, M Buch, N Choudhury, D Faluyi, T Felton, T Gorsuch, N A Hanley, T Hussell, Z Kausar, N Odell, R Osbourne, K Piper Hanley, K Radhakrishnan, S Stockdale.

**Newcastle upon Tyne Hospitals NHS Foundation Trust & University of Newcastle**

A De Soyza (PI), C Echevarria (PI), A Ayoub, J Brown, G Burns, G Davies, H Fisher, C Francis, A Greenhalgh, P Hogarth, J Hughes, K Jiwa, G Jones, G MacGowan, D Price, A Sayer, J Simpson, H Tedd, S Thomas, S West, M Witham, S Wright, A Young.

**NHS Dumfries and Galloway**

M J McMahon (PI), P Neill.

**NHS Greater Glasgow and Clyde Health Board & University of Glasgow**

D Anderson (PI), H Bayes (PI), C Berry (PI), D Grieve (PI), I B McInnes (PI), N Basu, A Brown, A Dougherty, K Fallon, L Gilmour, K Mangion, A Morrow, K Scott, R Sykes.

**NHS Highland**

E K Sage (PI), F Barrett, A Donaldson.

**NHS Lanarkshire**

M Patel (PI), D Bell, A Brown, M Brown, R Hamil, K Leitch, L Macliver, J Quigley, A Smith, B Welsh.

**NHS Lothian & University of Edinburgh**

G Choudhury (PI), J K Baillie, S Clohisey, A Deans, A B Docherty, J Furniss, E M Harrison, S Kelly, N I Lone, A Sheikh.

**NHS Tayside & University of Dundee**

J D Chalmers (PI), D Connell, A Elliott, C Deas, J George, S Mohammed, J Rowland, A R Solstice, D Sutherland, C J Tee.

**North Bristol NHS Trust & University of Bristol**

N Maskell (PI), D Arnold, S Barrett, H Adamali, A Dipper, S Dunn, A Morley, L Morrison, L Stadon, S Waterson, H Welch.

**North Middlesex Hospital NHS Trust**

B Jayaraman (PI), T Light.

**Nottingham University Hospitals NHS Trust & University of Nottingham**

C E Bolton (PI), P Almeida, J Bonnington, M Chrystal, C Dupont, P Greenhaff, A Gupta, L Howard, W Jang, S Linford, L Matthews, R Needham, A Nikolaidis, S Prosper, K Shaw, A K Thomas.

**Oxford University Hospitals NHS Foundation Trust & University of Oxford**

L P Ho (PI), N M Rahman (PI), M Ainsworth, A Alamoudi, A Bates, A Bloss, A Burns, P Carter, J Chen, F Conneh, T Dong, R I Evans, E Fraser, X Fu, J R Geddes, F Gleeson, P Harrison, M Havinden-Williams, P Jezzard, N Kanellakis, I Koychev, P Kurupati, X Li, H McShane, C Megson, K Motohashi, S Neubauer, D Nicoll, G Ogg, E Pacpaco, M Pavlides, Y Peng, N Petousi, N Rahman, B Raman, M J Rowland, K Saunders, M Sharpe, N Talbot, E Tunnicliffe.

**Royal Brompton and Harefield Clinical Group, Guy’s and St Thomas’ NHS Foundation Trust.**

W D-C Man (PI), B Patel (PI), R E Barker, D Cristiano, N Dormand, M Gummadi, S Kon, K Liyanage, C M Nolan, S Patel, O Polgar, P Shah, S J Singh, J A Walsh.

**Royal Free London NHS Foundation Trust**

J R Hurst (PI), H Jarvis (PI), S Mandal (PI), S Ahmad, S Brill, L Lim, D Matila, O Olaosebikan, C Singh.

**Royal Papworth Hospital NHS Foundation Trust**

M Toshner (PI), H Baxendale, L Garner, C Johnson, J Mackie, A Michael, J Pack, K Paques, H Parfrey, J Parmar.

**Salford Royal NHS Foundation Trust**

N Diar Bakerly (PI), P Dark, D Evans, E Hardy, A Harvey, D Holgate, S Knight, N Mairs, N Majeed, L McMorrow, J Oxton, J Pendlebury, C Summersgill, R Ugwuoke, S Whittaker.

**Salisbury NHS Foundation Trust**

W Matimba-Mupaya (PI), S Strong-Sheldrake.

**Sheffield Teaching NHS Foundation Trust & University of Sheffield**

S L Rowland-Jones (PI), A A R Thompson (Co PI), J Bagshaw, M Begum, K Birchall, R Butcher, H Carborn, F Chan, K Chapman, Y Cheng, L Chetham, C Clark, Z Coburn, J Cole, M Dixon, A Fairman, J Finnigan, H Foot, D Foote, A Ford, R Gregory, K Harrington, L Haslam, L Hesselden, J Hockridge, A Holbourn, B Holroyd-Hind, L Holt, A Howell, E Hurditch, F Ilyas, C Jarman, A Lawrie, E Lee, J-H Lee, R Lenagh, A Lye, I Macharia, M Marshall, A Mbuyisa, J McNeill, S Megson, J Meiring, L Milner, S Misra, H Newell, T Newman, C Norman, L Nwafor, D Pattenadk, M Plowright, J Porter, P Ravencroft, C Roddis, J Rodger, P Saunders, J Sidebottom, J Smith, L Smith, N Steele, G Stephens, R Stimpson, B Thamu, N Tinker, K Turner, H Turton, P Wade, S Walker, J Watson, I Wilson, A Zawia.

**St George’s University Hospitals NHS Foundation Trust**

R Aul (PI), M Ali, A Dunleavy (PI), D Forton, N Msimanga, M Mencias, T Samakomva, S Siddique, J Teixeira, V Tavoukjian.

**Sherwood Forest Hospitals NHS Foundation Trust**

J Hutchinson (PI), L Allsop, K Bennett, P Buckley, M Flynn, M Gill, C Goodwin, M Greatorex, H Gregory, C Heeley, L Holloway, M Holmes, J Kirk, W Lovegrove, TA Sewell, S Shelton, D Sissons, K Slack, S Smith, D Sowter, S Turner, V Whitworth, I Wynter.

**Shropshire Community Health NHS Trust**

L Warburton (PI), S Painter, J Tomlinson.

**Somerset NHS Foundation Trust**

C Vickers (PI), T Wainwright, D Redwood, J Tilley, S Palmer.

**Swansea Bay University Health Board**

G A Davies (PI), L Connor, A Cook, T Rees, F Thaivalappil, C Thomas.

**Tameside and Glossop Integrated Care NHS Foundation**

A Butt (PI), M Coulding, H Jones, S Kilroy, J McCormick, J McIntosh, H Savill, V Turner, J Vere.

**The Great Western Hospital Foundation Trust**

E Fraile (PI), J Ugoji.

**The Hillingdon Hospitals NHS Foundation Trust**

S S Kon (PI), H Lota, G Landers, M Nasseri, S Portukhay.

**The Rotherham NHS Foundation Trust**

A Hormis (PI), A Daniels, J Ingham, L Zeidan.

**United Lincolnshire Hospitals NHS Trust**

M Chablani (PI), L Osborne.

**University College London Hospital & University College London**

M Marks (PI), J S Brown (PI), N Ahwireng, B Bang, D Basire, R C Chambers, A Checkley, R Evans, M Heightman, T Hillman, J Hurst, J Jacob, S Janes, R Jastrub, M Lipman, S Logan, D Lomas, M Merida Morillas, H Plant, J C Porter, K Roy, E Wall.

**University Hospital Birmingham NHS Foundation Trust & University of Birmingham**

D Parekh (PI), N Ahmad Haider, C Atkin, R Baggott, M Bates, A Botkai, A Casey, B Cooper, J Dasgin, K Draxlbauer, N Gautam, J Hazeldine, T Hiwot, S Holden, K Isaacs, T Jackson, S Johnson, V Kamwa, D Lewis, J M Lord, S Madathil, C McGhee, K Mcgee, A Neal, A Newton Cox, J Nyaboko, D Parekh, Z Peterkin, H Qureshi, L Ratcliffe, E Sapey, J Short, T Soulsby, J Stockley, Z Suleiman, T Thompson, M Ventura, S Walder, C Welch, D Wilson, S Yasmin, K P Yip.

**University Hospitals of Derby and Burton**

P Beckett (PI) C Dickens, U Nanda.

**University Hospitals of Leicester NHS Trust & University of Leicester**

C E Brightling (CI), R A Evans (PI), M Aljaroof, N Armstrong, H Arnold, H Aung, M Bakali, M Bakau, M Baldwin, M Bingham, M Bourne, C Bourne, N Brunskill, P Cairns, L Carr, A Charalambou, C Christie, M J Davies, S Diver, S Edwards, C Edwardson, O Elneima, H Evans, J Finch, S Glover, N Goodman, B Gootpu, N J Greening, K Hadley, P Haldar, B Hargadon, V C Harris, L Houchen-Wolloff, W Ibrahim, L Ingram, K Khunti, A Lea, D Lee, G P McCann, H J C McAuley, P McCourt, T Mcnally, A Moss, W Monteiro, M Pareek, S Parker, A Rowland, A Prickett, I N Qureshi, R Russell, M Sereno, A Shikotra, S Siddiqui, A Singapuri, S J Singh, J Skeemer, M Soares, E Stringer, T Thornton, M Tobin, L V Wain, T J C Ward, F Woodhead, T Yates, A Yousuf.

**University Hospital Southampton NHS Foundation Trust & University of Southampton**

M G Jones (PI), C Childs, R Djukanovic, S Fletcher, M Harvey, E Marouzet, B Marshall, R Samuel, T Sass, T Wallis, H Wheeler.

**Whittington Health NHS**

R Dharmagunawardena (PI), E Bright, P Crisp, M Stern.

**Wirral University Teaching Hospital**

A Wight (PI), L Bailey, A Reddington.

**Wrightington Wigan and Leigh NHS trust**

A Ashish (PI), J Cooper, E Robinson.

**Yeovil District Hospital NHS Foundation Trust**

A Broadley (PI).

**York & Scarborough NHS Foundation Trust**

K Howard (PI), L Barman, C Brookes, K Elliott. L Griffiths, Z Guy, D Ionita, H Redfearn, C Sarginson, A Turnbull.

**Health and Care Research Wales**

Y Ellis.

**London School of Hygiene & Tropical Medicine (LSHTM)**

M Marks, A Briggs.

**NIHR Office for Clinical Research Infrastructure**

K Holmes.

**Patient Public Involvement Leads**

Asthma and Lung UK: K Poinasamy, S Walker.

**Royal Surrey NHS Foundation Trust**

M Halling-Brown.

**South London and Maudsley NHS Foundation Trust & Kings College London**

G Breen, M Hotopf.

**Swansea University & Swansea Welsh Network**

K Lewis, N Williams.

