## Supplement files for "Cohort Profile: Post-hospitalisation COVID-19 study (PHOSP-COVID)"

*PHOSP-COVID Collaborative Group*

### PHOSP-COVID Collaborative Group

**Core Management Group**

*Chief Investigator* C E Brightling, *Members* R A Evans (Lead Co-I), L V Wain (Lead Co-I), J D Chalmers, V C Harris, L P Ho, A Horsley, M Marks, K Poinasamy, B Raman, A Shikotra, A Singapuri

**PHOSP-COVID Study Central Coordinating Team**

C E Brightling (Chief Investigator), R A Evans (*Lead Co-I*), L V Wain (*Lead Co-I*), R Dowling, C Edwardson, O Elneima, S Finney, N J Greening, B Hargadon, V Harris, L Houchen--Wolloff, O C Leavy, H J C McAuley, C Overton, T Plekhanova, R M Saunders, M Sereno, A Singapuri, A Shikotra, C Taylor, S Terry, C Tong, B Zhao

**Steering Committee**

*Co-chairs* D Lomas, E Sapey*, Institution representatives* C Berry, C E Bolton, N Brunskill, E R Chilvers, R Djukanovic, Y Ellis, D Forton, N French, J George, N A Hanley, N Hart, L McGarvey, N Maskell, H McShane, M Parkes, D Peckham, P Pfeffer, A Sayer, A Sheikh, A A R Thompson, N Williams and core management group representation

**Executive Board**

*Chair* C E Brightling, representation from the core management group, each working group and platforms

**Platforms**

**Bioresource**

W Greenhalf (*Co-Lead*), M G Semple (*Co-Lead*), M Ashworth, H E Hardwick, L Lavelle-Langham, W Reynolds, M Sereno, R M Saunders, A Singapuri, V Shaw, A Shikotra, B Venson, L V Wain

M Marks, A Briggs

**NIHR Office for Clinical Research Infrastructure**

K Holmes

**Patient Public Involvement Leads**

Asthma UK and British Lung Foundation Partnership - K Poinasamy, S Walker

**Royal Surrey NHS Foundation Trust**

M Halling-Brown

**South London and Maudsley NHS Foundation Trust & Kings College London**

G Breen, M Hotopf

**Swansea University & Swansea Welsh Network**

K Lewis, N Williams

### Supplementary Methods

**Table SM1. Methods and thresholds for processing of variables and outcome measures presented in this analysis.**

|  | **Method** |
| --- | --- |
| **Tables 1 & S2** |  |
| Indices of Multiple Deprivation (IMD) | Obtained using postcode^1^. Different modalities used for the devolved nations. |
| Comorbidities | A pre-existing comorbidity was considered absent if not indicated by a ‘yes’ on the case report form. |
| Admission duration | Calculated using the hospital discharge date and the earliest admission date to the same or different hospital for the participant’s COVID-19 episode. |
| WHO clinical progression scale | WHO classes are as follows: 3–4=no continuous supplemental oxygen needed; 5=continuous supplemental oxygen only; 6=continuous or bi-level positive airway pressure ventilation or high-flow nasal oxygen; and 7–9=invasive mechanical ventilation or other organ support e.g. vasopressors, dialysis or Extracorporeal membrane oxygenation (ECMO) ^2^. |
| **Table 3** |  |
| Generalised Anxiety Disorder Questionnaire (GAD-7) (Anxiety) | The Generalised Anxiety Disorder (GAD-7) questionnaire is a patient reported outcome measure consisting of 7 questions with total scores ranging from 0 to 21. We used a GAD7 threshold score of > 8 to suggest at least mild-moderate anxiety.^3^ |
| Patient Health Questionnaire (PHQ-9) (Depression) | The Patient Health Questionnaire (PHQ-9) is a patient reported outcome measure consisting of 9 questions with total scores ranging from 0 to 27. We used a PHQ-9 threshold score of ≥10 to suggest at least moderate depression.^4^ |
| Post-Traumatic Stress Disorder Checklist for DSM V (PCL-5) Questionnaire | The Post-Traumatic Stress Disorder Checklist for DSM V (PCL-5) questionnaire is a patient reported outcome measure consisting of 20 questions assessing evidence of post-traumatic stress disorder according to the DSM V criteria. Total scores range from 0-80. We used a PCL-5 threshold score of ≥38 suggestive of a provisional diagnosis of post-traumatic stress disorder.^5,6^ |
| Dyspnoea-12 | The Dyspnoea-12 questionnaire is a patient reported outcome measure consisting of 12 questions assessing breathlessness severity incorporating both “physical” and “affective” aspects^7^. Scores range from 0 to 36 with higher scores corresponding to greater severity of breathlessness. |
| FACIT fatigue subscale score (FACIT) | The Functional Assessment of Chronic Illness Therapy – Fatigue (FACIT-Fatigue) scale is a patient reported outcome measure consisting of 13 questions to assess self-reported fatigue and its impact on daily activities and function.^8^ Total scores range from 0-52, with lower scores corresponding to an increased burden of fatigue.^9^ |
| Brief Pain Inventory (BPI) severity and interference | The Brief Pain Inventory (BPI) is a patient reported outcome questionnaire consisting of 15 questions across domains of pain severity and pain interference. We have reported the BPI Severity score as the mean score from the 4 severity questions each with a range 0 – 10 anchored at 0 = “No Pain” and 10 = “Pain as bad as you can imagine”.^10,11^ |
| Short Physical Performance Battery (SPPB) | The Short Physical Performance Battery (SPPB) test is a researcher administered assessment of physical performance and frailty. It comprises 3 components; balance, gait speed and sit to stand tests. Tests were completed according to recommended standards and training was provided to site staff by the central study team via a recorded demonstration video. SPPB total scores range from 0-12. We have reported a total SPPB score of ≤10 suggestive of underlying frailty.^12-14^ |
| Incremental Shuttle Walk Test (ISWT) | The Incremental Shuttle Walk Test (ISWT) is a researcher administered assessment of maximal physical performance and was performed according to standardised instructions with two attempts performed by participants on the same day with a 20 minutes rest between them.^15^ Training was provided to site staff by the central study team via a recorded demonstration video. The best effort was reported in metres and the percent predicted value was calculated using the following reference formula accounting for gender, age and BMI.^16^ (ISWT predicted = 1449·701 − (11·735 × age) + (241·897 × gender) − (5·686 × BMI), where male gender = 1 and female gender = 0) |
| Rockwood Clinical Frailty Scale (CFS) | The Rockwood Clinical Frailty Scale (CFS) is a researcher assessed scale of clinical frailty with scores ranging from 1-9 where lower scores correspond to increased frailty. We have reported CFS scores of <5 suggestive of frailty.^17^ |
| Montreal Cognitive Assessment (MoCA) | The Montreal Cognitive Assessment (MoCA) is a researcher administered cognitive function questionnaire across 8 domains. Training was provided to site staff using standardised resources supplied online by MoCA TEST Inc.^18^ The assessment was conducted in English with researchers applying their discretion to exclude participants whose command of English was insufficient to complete the test accurately. Total scores range from 0 to 30. We report total MoCA scores of <23 suggestive of at least Mild Cognitive Impairment.^19^ |
| Spirometry and Pulmonary Function Testing | Due to COVID-19 related restrictions on aerosol-generating procedures during the study period, access to spirometry and lung function was limited. Spirometry and Pulmonary function testing were completed as per ERS/ATS recommendations.^20^ Spirometry and Transfer factor values were converted to SI units if not reported as such by sites. Transfer Capacity of the Lung for the uptake of carbon monoxide (TLCO) and carbon monoxide transfer coefficient (KCO) were obtained from the best of two repeat readings. ERS Reference values were used to calculate % predicted values.^21-23^ FEV_1_/FVC <0·7 was used to define airflow obstruction.^24^ % predicted TLCO <80% was considered indicative of impaired gas transfer. |
| BNP / NT-pro BNP | Brain Natriuretic Peptide (BNP) or N-terminal pro B-type Natriuretic Peptide (NT-pro BNP) were collected according to each site’s routine clinically available assay as a biomarker of heart failure. Three sites submitted BNP results with all of the remaining sites submitting NT-pro BNP results. The threshold values used for BNP was ≥ 100 ng/litre ^25^ and for NT-pro BNP ≥ 400ng/litre ^26^ as suggestive of heart failure. |
| Glycated haemoglobin (HbA1c) | Glycated haemoglobin (HbA1c) was collected as a biomarker of current glycaemic control. We have reported HbA1c levels ≥ 6·5% as suggestive of a diagnosis of diabetes.^27^ |
| C-Reactive Protein (CRP) | C-Reactive Protein (CRP) levels were collected as a biomarker of current systemic inflammation. Values reported as below the lower or upper limit reportable range for the assay used at the site have been included at the stated less than or more than cut off value for calculation of mean (SD) results. We have reported CRP levels > 5 mg/L as suggestive of systemic inflammation. |
| **Table 4** |  |
| Symptoms at five-month and one-year visits | Symptom severity was rated using a 0-10 visual analogue scale for Breathlessness, Cough, Fatigue, Sleep quality and Pain before COVID-19 illness and worst in the last 24 hours. |
| EQ-5D-5L VAS | The EQ5D Visual Analogue Scale is a patient reported outcome questionnaire recording the patient’s self-rated health and was completed for “before your COVID-19 illness” and “your own health state today.” Scores are presented as mean and standard deviation.^28^ |
| EQ-5D-5L Utility Index | The EQ5D-5L is a five-dimension patient reported outcome questionnaire recording a patient’s self-rated health state for mobility, self-care, usual activities, pain/discomfort and anxiety/depression. These scores are then mapped to a United Kingdom specific Utility Index anchored at 1 for “perfect health” and 0 for “dead” calculated from reported EQ5D-5L scores across the five dimensions.^29^ |
| Washington Group Short Set of Functioning Severity Continuum | The Washington Group Short Set of Functioning (WG-SS) is a patient reported outcome questionnaire using six questions to assess disability and function. Participant responses were transformed to the “Severity Continuum” by assigning scores of zero to responses “no difficulty”, one to responses “some difficulty”, six to responses “a lot of difficulty” and 36 to responses “cannot do at all”. ^30^ |

### Supplementary Data – Results

**Table S1. The participants’ pre-existing comorbidities stratified by tier allocation.**

|  | **Complete PHOSP cohort (n=7935)** | **Tier 1**  **(n=5238)** | **Tier 2**  **(n=2697)** |
| --- | --- | --- | --- |
| **Cardiovascular** |  |  |  |
| Myocardial Infarction | 375 (4.7%) | 259 (4.9%) | 116 (4.3%) |
| Ischaemic Heart Disease | 535 (4.67%) | 370 (7.1%) | 165 (6.1%) |
| Atrial Fibrillation | 473 (5.9%) | 348 (6.6%) | 125 (4.6%) |
| Hypertension | 2946 (37.1%) | 2012 (38.4%) | 934 (34.6%) |
| Congestive Heart Failure | 169 (2.1%) | 132 (2.5%) | 37 (1.4%) |
| Congenital heart disease | 38 (0.5%) | 26 (0.5%) | 12 (0.4%) |
| Valvular heart disease | 115 (1.4%) | 80 (1.5%) | 35 (1.3%) |
| Pacemaker/Implantable defibrillator | 106 (1.3%) | 83 (1.6%) | 23 (0.9%) |
| Peripheral Vascular Disease | 104 (1.3%) | 67 (1.3%) | 37 (1.4%) |
| Hypercholesterolemia/dyslipidaemia | 1,289 (16.2%) | 771 (14.7%) | 518 (19.2%) |
| Cerebrovascular Accident/ Transient  Ischaemic Attack | 304 (3.8%) | 195 (3.7%) | 109 (4.0%) |
| **Neurological and Psychiatric** |  |  |  |
| Dementia | 27 (0.3%) | 22 (0.4%) | 5 (0.2%) |
| Anxiety or Depression | 1367 (17.2%) | 912 (17.4%) | 455 (16.9%) |
| Chronic fatigue syndrome/ fibromyalgia or chronic pain | 339 (4.3%) | 199 (3.8%) | 140 (5.2%) |
| Previous treatment with antidepressants | 921 (11.6%) | 593 (11.3%) | 328 (12.2%) |
| Any previous treatment by a mental health professional | 349 (4.4%) | 191 (3.6%) | 158 (5.9%) |
| **Respiratory** |  |  |  |
| COPD | 602 (7.6%) | 461 (8.8%) | 141 (5.2%) |
| Asthma | 1481 (18.7%) | 999 (19.1%) | 482 (17.9%) |
| Interstitial lung disease | 37 (0.5%) | 28 (0.5%) | 9 (0.3%) |
| Bronchiectasis | 120 (1.5%) | 79 (1.5%) | 41 (1.5%) |
| Obstructive sleep apnoea | 362 (4.6%) | 226 (4.3%) | 136 (5.0%) |
| Obesity hypoventilation syndrome | 21 (0.3%) | 14 (0.3%) | 7 (0.3%) |
| Pleural effusion | 49 (0.6%) | 34 (0.6%) | 15 (0.6%) |
| **Rheumatological** |  |  |  |
| Connective tissue disease | 35 (0.4%) | 23 (0.4%) | 12 (0.4%) |
| Rheumatoid arthritis | 247 (3.1%) | 166 (3.2%) | 81 (3.0%) |
| Osteoarthritis | 813 (10.2%) | 545 (10.4%) | 268 (9.9%) |
| **Gastrointestinal** |  |  |  |
| Peptic ulcer disease | 80 (1.0%) | 49 (0.9%) | 31 (1.1%) |
| Liver Disease - *mild* | 98 (1.2%) | 57 (1.1%) | 41 (1.5%) |
| Liver Disease – *moderate/severe* | 76 (0.9%) | 39 (0.7%) | 37 (1.4%) |
| Gastro-oesophageal reflux disease | 665 (8.4%) | 379 (7.2%) | 286 (10.6%) |
| Inflammatory bowel disease | 116 (1.5%) | 72 (1.4%) | 44 (1.6%) |
| Irritable bowel syndrome | 211 (2.7%) | 132 (2.5%) | 79 (2.9%) |
| **Metabolic/Endocrine/Renal** |  |  |  |
| Type 1 diabetes | 79 (1.0%) | 51 (1.0%) | 28 (1.0%) |
| Type 2 diabetes | 1683 (21.2%) | 1146 (21.9%) | 537 (19.9%) |
| Hypothyroidism | 465 (5.9%) | 322 (6.1%) | 143 (5.3%) |
| Hyperthyroidism | 66 (0.8%) | 41 (0.8%) | 25 (0.9%) |
| Chronic Kidney Disease | 432 (5.4%) | 322 (6.1%) | 110 (4.1%) |
| **Malignancy** |  |  |  |
| Solid Tumour Malignancy - localised | 319 (4.0%) | 227 (4.3%) | 92 (3.4%) |
| Solid Tumour Malignancy - metastatic | 42 (0.5%) | 33 (0.6%) | 9 (0.3%) |
| Leukaemia | 53 (0.7%) | 31 (0.6%) | 22 (0.8%) |
| Lymphoma | 53 (0.7%) | 35 (0.7%) | 18 (0.7%) |
| **Chronic Infectious Diseases** |  |  |  |
| HIV | 22 (0.3%) | 14 (0.3%) | 8 (0.3%) |
| Chronic viral hepatitis | 57 (0.7%) | 30 (0.6%) | 27 (1.0%) |
| Mycobacterium Tuberculosis | 65 (0.8%) | 40 (0.8%) | 25 (0.9%) |

**Table S2: Comparison of the participants’ characteristics between the 1-year visit attendees and Tier 2 participants who failed to return to a 1-year visit.**

|  | **Participants who didn’t attend a 1-year visit (n=597)** | **Participants who attended a 1-year visit (n=2100)** |
| --- | --- | --- |
| Admission at age, years† | 54.6 (13.6) | 58.9 (12.1) |
| Sex |  |  |
| Female | 227 (38.1%) | 811 (38.6%) |
| Male | 369 (61.9%) | 1289 (61.4%) |
| Ethnicity |  |  |
| White | 398 (67.7%) | 1609 (76.9%) |
| South Asian | 100 (17.0%) | 205 (9.8%) |
| Black | 51 (8.7%) | 142 (6.8%) |
| Mixed | 10 (1.7%) | 45 (2.2%) |
| Other | 29 (4.9%) | 89 (4.3%) |
| Missing data | 9 | 10 |
| Index of multiple deprivation score (IMD) |  |  |
| 1 (most deprived) | 152 (25.9%) | 466 (22.3%) |
| 2 | 162 (27.7%) | 460 (22.0%) |
| 3 | 81 (13.8%) | 382 (18.3%) |
| 4 | 101 (17.2%) | 371 (17.7%) |
| 5 (least deprived) | 90 (15.4%) | 412 (19.7%) |
| Missing data | 11 | 9 |
| Body-mass index (BMI) |  |  |
| Median (IQR) †† | 31.1 [27.3 - 36.2] | 31.2 [27.7- 35.6] |
| <30 kg/m^2^ | 182 (45.0%) | 613 (41.7%) |
| ≥30 kg/m^2^ | 222 (55.0%) | 856 (58.3%) |
| Missing data | 193 | 631 |
| Healthcare worker | 97 (17.6%) | 279 (13.9%) |
| Missing data | 46 | 96 |
| WHO clinical progression scale |  |  |
| WHO class 3-4 | 126 (21.1%) | 321 (15.3%) |
| WHO class 5 | 240 (40.2%) | 895 (42.6%) |
| WHO class 5 | 129 (21.6%) | 504 (24.0%) |
| WHO class 7-9 | 102 (17.1%) | 380 (18.1%) |
| Comorbidities |  |  |
| Median number of comorbidities (IQR) | 1 [0-3] | 2 [1-3] |
| 0 | 172 (28.8%) | 495 (23.6%) |
| 1 | 129 (21.6%) | 442 (21.0%) |
| ≥2 | 296 (49.6%) | 1163 (55.4%) |
| Cardiovascular | 247 (41.4%) | 992 (47.2%) |
| Respiratory | 154 (25.8%) | 570 (27.1%) |
| Type 2 diabetes | 110 (18.5%) | 427 (20.4%) |
| Neuro-psychiatric | 131 (21.9%) | 431 (20.5%) |
| Renal and endocrine | 53 (8.9%) | 234 (11.1%) |
| Admission duration, days† | 12.1 (14.9) | 14.6 (18.7) |
| Positive SARS-CoV-2 PCR | 495 (92.4%) | 1788 (92.6%) |
| Missing data | 61 | 169 |
| Systemic steroids | 293 (51.7%) | 1,155 (57.9%) |
| Missing data | 30 | 106 |
| Antibiotic therapy | 468 (79.9%) | 1,607 (78.5%) |
| Missing data | 11 | 54 |
| Anti-coagulants | 245 (42.7%) | 928 (46.6%) |
| Missing data | 23 | 109 |
| Recovery cluster at 5-month visit | 524 |  |
| Very Severe | 117 (22.3%) | 386 (20.5%) |
| Severe | 134 (25.6%) | 502 (26.7%) |
| Moderate with cognitive impairment | 117 (22.3%) | 426 (22.7%) |
| Mild | 156 (29.8%) | 567 (30.1%) |
| Missing data | 73 | 219 |

Data are n (%) unless † mean (SD) or †† median [IQR]. Percentages are calculated by category after exclusion of missing data for that variable. WHO classes are as follows: 3–4=no continuous supplemental oxygen needed; 5=continuous supplemental oxygen only; 6=continuous or bi-level positive airway pressure ventilation or high-flow nasal oxygen; and 7–9=invasive mechanical ventilation or other organ support. IMD=Index of Multiple Deprivation. BMI=body-mass index. See Table S1 for further descriptions of variables.

**Table S3. Total number of received biological research samples.**

|  | **Time points** | | |
| --- | --- | --- | --- |
| **Kit Type** | **5-month** | **1-year** | **Total** |
| Blood | 2,308 | 1,779 | 4,087 |
| Oral rinse | 1,626 | 1,394 | 3,020 |
| Sputum (spontaneous) | 326 | 197 | 523 |
| Urine | 2,085 | 1,706 | 3,791 |
| Saliva (for DNA analysis) * | - | - | 2,638 |

*Saliva samples were collected once from a subset of tier 1 participants only.

**Table S4. Total number of acquired radiological images (X-rays and thoracic CT scans).**

| **Type of imaging** | **Tier 1** | **Tier 2** | **Total** |
| --- | --- | --- | --- |
| Non-contrast thoracic CT scans | 113 | 260 | 373 |
| Contrast thoracic CT scans | 93 | 207 | 300 |
| Chest X-Rays | - | - | 3903 |

**Table S5. Total number of acquired MRI scans as per the C-MORE sub-study per contributing sites.**

| **Centre Name** | **N of participants** | **Received Studies** |
| --- | --- | --- |
| University Hospitals of Leicester NHS Trust | 99 | 99 |
| Oxford University Hospitals NHS Foundation Trust | 61 | 61 |
| Sheffield Teaching Hospitals NHS Foundation Trust | 48 | 48 |
| Leeds Teaching Hospitals NHS Trust | 39 | 47 |
| Imperial College Healthcare NHS Trust | 34 | 34 |
| Kings College Hospital NHS Foundation Trust | 33 | 33 |
| University College London Hospitals NHS Foundation Trust | 33 | 45 |
| Manchester University NHS Foundation Trust | 24 | 24 |
| Liverpool University Hospitals NHS Foundation Trust | 20 | 42 |
| Nottingham University Hospitals NHS Trust | 13 | 13 |
| St Bartholomews Hospital, Barts Health NHS Trust | 10 | 11 |
| University Hospitals Birmingham NHS Foundation Trust | 10 | 47 |
| Royal Papworth Hospital | 8 | 7 |
| Total | 432 | 511 |

**Table S6. Final list of top 10 research questions (not ranked).**

| **No** | **Research question** |
| --- | --- |
| 1. | What are the underlying mechanisms of long COVID that drive symptoms and/or organ impairment? |
| 2. | What imaging techniques or scans may be able to detect and predict the development of organ problems or wider systemic issues? |
| 3. | What happens to the immune system throughout patients’ recovery from COVID-19? |
| 4. | What can data at 6 and 12 months tell us about the long-term trajectory of illness? |
| 5. | What blood or other laboratory tests may be able to detect and predict the development of organ problems or wider systemic issues? |
| 6. | What is the impact of treatment(s) during the acute (initial) stage of COVID-19 on recovery? |
| 7. | What are the problems within the muscles associated with symptoms limiting activity/function/exercise? If so, what can be done to help? |
| 8. | What medications, dietary changes, supplements, rehabilitation and therapies aid recovery? |
| 9. | What can be done to support mental well-being during recovery? |
| 10. | What is the risk of future adverse health events (e.g., stroke, heart attack)? |

**Table S7. Change in occupation status COVID stratified by the two research visits.**

|  | **5-month (n=2570)** | | **1-year (n=2100)** | |
| --- | --- | --- | --- | --- |
|  | **n** | **%** | **n** | **%** |
| Working full-time or part-time before COVID-19 | 1137 | 54.2% | 914 | 52.8% |
| Missing data | 471 |  | 369 |  |
| No longer working after COVID-19 | 222 | 11.0% | 121 | 8.5% |
| Missing data | 554 |  | 681 |  |
| Occupation change due to health after COVID-19 | 803 | 46.7% | 326 | 34.6% |
| Missing data | 849 |  | 1159 |  |

### Appendix 1 - Patient Symptom Questionnaire (PSQ)

| **Patient Symptom Questionnaire** *(Participants to complete)* | | |
| --- | --- | --- |
| **Post-hospitalisation COVID-19 Follow-Up** | | |
| Date of completion |  | **Patient ID sticker or**  **Name: _____________________________**  **Date of birth: ____ / _______ / ________**  *(if photocopied for research purposes, please redact participant identifiable information)* |
| **☐ ☐ / ☐ ☐ ☐ / ☐☐☐☐**  d d m m m y y y y | |  |

| **Please complete the following questionnaire**  If anything is unclear or there is something you are concerned about, please discuss this with your clinician or nurse during your appointment. | | | | | | | | | | | | | | | | | | | | | | | | | | | | | | | | | | | | | | | | | | | | | | | | | | | | | | | |
| --- | --- | --- | --- | --- | --- | --- | --- | --- | --- | --- | --- | --- | --- | --- | --- | --- | --- | --- | --- | --- | --- | --- | --- | --- | --- | --- | --- | --- | --- | --- | --- | --- | --- | --- | --- | --- | --- | --- | --- | --- | --- | --- | --- | --- | --- | --- | --- | --- | --- | --- | --- | --- | --- | --- | --- |
| a) Do you feel fully recovered from COVID-19? | | | | | | | | | | | | | | | | | | | | | | | | | | |  | | | | | | | | | | | | | |  | | | | | | | | |  | | | | | |
| **☐ Yes** | | | | | | | | | | | **☐ No** | | | | | | | | | | | | | | | | | | | | | | | | | | | | | **☐ Not sure** | | | | | | | | | | | | | | | |
| **We would like to understand more about your symptoms. For each symptom below (b to f), please rate your symptoms for how you are now (since you had COVID-19) and the worst you have felt in the last 24 hours.**  **Please rate these symptoms on a scale of 0 – 10**  **Please also try to indicate if the symptom is staying the same, getting better or getting worse (trajectory)** | | | | | | | | | | | | | | | | | | | | | | | | | | | | | | | | | | | | | | | | | | | | | | | | | | | | | | | |
|  | | |  | | ← | | | **0** | | | | | | **1** | **2** | | | | **3** | | | | | | | **4** | | | | **5** | | | | **6** | | | | | | | | | **7** | | | **8** | | | | | | **9** | | | **10** |
|  | | |  | | NEUTRAL | | | | | | | | | SLIGHT SENSATION | | | | | | | |  | | | | ANNOYING | | | | | | | | DISTRESSING | | | | | | | | | | | | | | | | | | UNBEARABLE | | | |
| b) Breathlessness | | | How you are now | | | | | **0** | | | | | | **1** | **2** | | | | **3** | | | | | | | **4** | | | | **5** | | | | **6** | | | | | | | | | **7** | | | **8** | | | | | | **9** | | | **10** |
|  |  |  |  |  |  |  |  | **☐** | | | | | | **☐** | **☐** | | | | **☐** | | | | | | | **☐** | | | | **☐** | | | | **☐** | | | | | | | | | **☐** | | | **☐** | | | | | | **☐** | | | **☐** |
|  |  |  | Worst in last 24hrs | | | | | **0** | | | | | | **1** | **2** | | | | **3** | | | | | | | **4** | | | | **5** | | | | **6** | | | | | | | | | **7** | | | **8** | | | | | | **9** | | | **10** |
|  |  |  |  |  |  |  |  | **☐** | | | | | | **☐** | **☐** | | | | **☐** | | | | | | | **☐** | | | | **☐** | | | | **☐** | | | | | | | | | **☐** | | | **☐** | | | | | | **☐** | | | **☐** |
|  |  |  | Trajectory | | | | | **☐ Same ☐ Better ☐ Worse** | | | | | | | | | | | | | | | | | | | | | | | | | | | | | | | | | | | | | | | | | | | | | | | |
| c) Cough | | | How you are now | | | | | **0** | | | | | | **1** | **2** | | | | **3** | | | | | | | **4** | | | | **5** | | | | **6** | | | | | | | | | **7** | | | **8** | | | | | | **9** | | | **10** |
|  |  |  |  |  |  |  |  | **☐** | | | | | | **☐** | **☐** | | | | **☐** | | | | | | | **☐** | | | | **☐** | | | | **☐** | | | | | | | | | **☐** | | | **☐** | | | | | | **☐** | | | **☐** |
|  |  |  | Worst in last 24hrs | | | | | **0** | | | | | | **1** | **2** | | | | **3** | | | | | | | **4** | | | | **5** | | | | **6** | | | | | | | | | **7** | | | **8** | | | | | | **9** | | | **10** |
|  |  |  |  |  |  |  |  | **☐** | | | | | | **☐** | **☐** | | | | **☐** | | | | | | | **☐** | | | | **☐** | | | | **☐** | | | | | | | | | **☐** | | | **☐** | | | | | | **☐** | | | **☐** |
|  |  |  | Trajectory | | | | | **☐ Same ☐ Better ☐ Worse** | | | | | | | | | | | | | | | | | | | | | | | | | | | | | | | | | | | | | | | | | | | | | | | |
| d) Fatigue | | | How you are now | | | | | **0** | | | | | | **1** | **2** | | | | **3** | | | | | | | **4** | | | | **5** | | | | **6** | | | | | | | | | **7** | | | **8** | | | | | | **9** | | | **10** |
|  |  |  |  |  |  |  |  | **☐** | | | | | | **☐** | **☐** | | | | **☐** | | | | | | | **☐** | | | | **☐** | | | | **☐** | | | | | | | | | **☐** | | | **☐** | | | | | | **☐** | | | **☐** |
|  |  |  | Worst in last 24hrs | | | | | **0** | | | | | | **1** | **2** | | | | **3** | | | | | | | **4** | | | | **5** | | | | **6** | | | | | | | | | **7** | | | **8** | | | | | | **9** | | | **10** |
|  |  |  |  |  |  |  |  | **☐** | | | | | | **☐** | **☐** | | | | **☐** | | | | | | | **☐** | | | | **☐** | | | | **☐** | | | | | | | | | **☐** | | | **☐** | | | | | | **☐** | | | **☐** |
|  |  |  | Trajectory | | | | | **☐ Same ☐ Better ☐ Worse** | | | | | | | | | | | | | | | | | | | | | | | | | | | | | | | | | | | | | | | | | | | | | | | |
| e) Sleep quality | | | How you are now | | | | | **0** | | | | | | **1** | **2** | | | | **3** | | | | | | | **4** | | | | **5** | | | | **6** | | | | | | | | | **7** | | | **8** | | | | | | **9** | | | **10** |
|  |  |  |  |  |  |  |  | **☐** | | | | | | **☐** | **☐** | | | | **☐** | | | | | | | **☐** | | | | **☐** | | | | **☐** | | | | | | | | | **☐** | | | **☐** | | | | | | **☐** | | | **☐** |
|  |  |  | Worst in last 24hrs | | | | | **0** | | | | | | **1** | **2** | | | | **3** | | | | | | | **4** | | | | **5** | | | | **6** | | | | | | | | | **7** | | | **8** | | | | | | **9** | | | **10** |
|  |  |  |  |  |  |  |  | **☐** | | | | | | **☐** | **☐** | | | | **☐** | | | | | | | **☐** | | | | **☐** | | | | **☐** | | | | | | | | | **☐** | | | **☐** | | | | | | **☐** | | | **☐** |
|  |  |  | Trajectory | | | | | **☐ Same ☐ Better ☐ Worse** | | | | | | | | | | | | | | | | | | | | | | | | | | | | | | | | | | | | | | | | | | | | | | | |
| f) Pain | | | How you are now | | | | | **0** | | | | | | **1** | **2** | | | | **3** | | | | | | | **4** | | | | **5** | | | | **6** | | | | | | | | | **7** | | | **8** | | | | | | **9** | | | **10** |
|  |  |  |  |  |  |  |  | **☐** | | | | | | **☐** | **☐** | | | | **☐** | | | | | | | **☐** | | | | **☐** | | | | **☐** | | | | | | | | | **☐** | | | **☐** | | | | | | **☐** | | | **☐** |
|  |  |  | Worst in last 24hrs | | | | | **0** | | | | | | **1** | **2** | | | | **3** | | | | | | | **4** | | | | **5** | | | | **6** | | | | | | | | | **7** | | | **8** | | | | | | **9** | | | **10** |
|  |  |  |  |  |  |  |  | **☐** | | | | | | **☐** | **☐** | | | | **☐** | | | | | | | **☐** | | | | **☐** | | | | **☐** | | | | | | | | | **☐** | | | **☐** | | | | | | **☐** | | | **☐** |
|  |  |  | Trajectory | | | | | **☐ Same ☐ Better ☐ Worse** | | | | | | | | | | | | | | | | | | | | | | | | | | | | | | | | | | | | | | | | | | | | | | | |
| g) How many hours sleep do you get in general every 24 hours? (to the nearest hour) | | | | | | | | | | | | | | | | | | | | | | | | | | | | | | | | | **☐ ☐ hours** | | | | | | | | | | | | | | | | | | | | | | |
| h) Within the **last seven days**, have you had any of these symptoms? | | | | | | | | | | | | | | | | | | | | | | | | | | | | | | | | | | | | | | | | | | | | | | | | | | | | | | | |
| 1. Breathlessness | | | | | | | | | | | | | | | | | | | | | | | | | | | | | | | | | ☐ Yes ☐ No | | | | | | | | | | | | | | | | | | | | | | |
| 2. Cough | | | | | | | | | | | | | | | | | | | | | | | | | | | | | | | | | ☐ Yes ☐ No | | | | | | | | | | | | | | | | | | | | | | |
| 3. Fatigue | | | | | | | | | | | | | | | | | | | | | | | | | | | | | | | | | ☐ Yes ☐ No | | | | | | | | | | | | | | | | | | | | | | |
| 4. Poor sleep | | | | | | | | | | | | | | | | | | | | | | | | | | | | | | | | | ☐ Yes ☐ No | | | | | | | | | | | | | | | | | | | | | | |
| 5. Pain | | | | | | | | | | | | | | | | | | | | | | | | | | | | | | | | | ☐ Yes ☐ No | | | | | | | | | | | | | | | | | | | | | | |
| **Neurological and other** | | | | | | | | | | | | | | | | | | | | | | | | | | | | | | | | | | | | | | | | | | | | | | | | | | | | | | | |
| 6. Loss of sense of smell | | | | | | | | | | | | | | | | | | | | | | | | | | | | | | | | | ☐ Yes ☐ No | | | | | | | | | | | | | | | | | | | | | | |
| 7. Loss of taste | | | | | | | | | | | | | | | | | | | | | | | | | | | | | | | | | ☐ Yes ☐ No | | | | | | | | | | | | | | | | | | | | | | |
| 8. Confusion/fuzzy head | | | | | | | | | | | | | | | | | | | | | | | | | | | | | | | | | ☐ Yes ☐ No | | | | | | | | | | | | | | | | | | | | | | |
| 9. Difficulty with communication | | | | | | | | | | | | | | | | | | | | | | | | | | | | | | | | | ☐ Yes ☐ No | | | | | | | | | | | | | | | | | | | | | | |
| 10. Difficulty with concentration | | | | | | | | | | | | | | | | | | | | | | | | | | | | | | | | | ☐ Yes ☐ No | | | | | | | | | | | | | | | | | | | | | | |
| 11. Short term memory loss | | | | | | | | | | | | | | | | | | | | | | | | | | | | | | | | | ☐ Yes ☐ No | | | | | | | | | | | | | | | | | | | | | | |
| 12. Physical slowing down | | | | | | | | | | | | | | | | | | | | | | | | | | | | | | | | | ☐ Yes ☐ No | | | | | | | | | | | | | | | | | | | | | | |
| 13. Slowing down in your thinking | | | | | | | | | | | | | | | | | | | | | | | | | | | | | | | | | ☐ Yes ☐ No | | | | | | | | | | | | | | | | | | | | | | |
| 14. Headache | | | | | | | | | | | | | | | | | | | | | | | | | | | | | | | | | ☐ Yes ☐ No | | | | | | | | | | | | | | | | | | | | | | |
| 15. Altered personality/behaviour (not the same person) | | | | | | | | | | | | | | | | | | | | | | | | | | | | | | | | | ☐ Yes ☐ No | | | | | | | | | | | | | | | | | | | | | | |
| 16. Limb weakness | | | | | | | | | | | | | | | | | | | | | | | | | | | | | | | | | ☐ Yes ☐ No | | | | | | | | | | | | | | | | | | | | | | |
| 17. Problems with balance | | | | | | | | | | | | | | | | | | | | | | | | | | | | | | | | | ☐ Yes ☐ No | | | | | | | | | | | | | | | | | | | | | | |
| 18. Can’t move and / or feel one side of your body or face | | | | | | | | | | | | | | | | | | | | | | | | | | | | | | | | | ☐ Yes ☐ No | | | | | | | | | | | | | | | | | | | | | | |
| 19. Problems seeing | | | | | | | | | | | | | | | | | | | | | | | | | | | | | | | | | ☐ Yes ☐ No | | | | | | | | | | | | | | | | | | | | | | |
| 20. Tingling feeling/“pins and needles“ | | | | | | | | | | | | | | | | | | | | | | | | | | | | | | | | | ☐ Yes ☐ No | | | | | | | | | | | | | | | | | | | | | | |
| 21. Can’t fully move or control movement | | | | | | | | | | | | | | | | | | | | | | | | | | | | | | | | | ☐ Yes ☐ No | | | | | | | | | | | | | | | | | | | | | | |
| 22. Tremor/shakiness | | | | | | | | | | | | | | | | | | | | | | | | | | | | | | | | | ☐ Yes ☐ No | | | | | | | | | | | | | | | | | | | | | | |
| 23. Seizures | | | | | | | | | | | | | | | | | | | | | | | | | | | | | | | | | ☐ Yes ☐ No | | | | | | | | | | | | | | | | | | | | | | |
| **Musculo-Skeletal** | | | | | | | | | | | | | | | | | | | | | | | | | | | | | | | | | | | | | | | | | | | | | | | | | | | | | | | |
| 24. Aching in your muscles (pain) | | | | | | | | | | | | | | | | | | | | | | | | | | | | | | | | | ☐ Yes ☐ No | | | | | | | | | | | | | | | | | | | | | | |
| 25. Joint pain or swelling | | | | | | | | | | | | | | | | | | | | | | | | | | | | | | | | | ☐ Yes ☐ No | | | | | | | | | | | | | | | | | | | | | | |
| **Cardio-Respiratory** | | | | | | | | | | | | | | | | | | | | | | | | | | | | | | | | | | | | | | | | | | | | | | | | | | | | | | | |
| 26. Leg/ankle swelling | | | | | | | | | | | | | | | | | | | | | | | | | | | | | | | | | ☐ Yes ☐ No | | | | | | | | | | | | | | | | | | | | | | |
| 27. Chest pain | | | | | | | | | | | | | | | | | | | | | | | | | | | | | | | | | ☐ Yes ☐ No | | | | | | | | | | | | | | | | | | | | | | |
| 28. Chest tightness | | | | | | | | | | | | | | | | | | | | | | | | | | | | | | | | | ☐ Yes ☐ No | | | | | | | | | | | | | | | | | | | | | | |
| 29. Pain on breathing | | | | | | | | | | | | | | | | | | | | | | | | | | | | | | | | | ☐ Yes ☐ No | | | | | | | | | | | | | | | | | | | | | | |
| 30. Palpitations | | | | | | | | | | | | | | | | | | | | | | | | | | | | | | | | | ☐ Yes ☐ No | | | | | | | | | | | | | | | | | | | | | | |
| 31. Dizziness or lightheadness | | | | | | | | | | | | | | | | | | | | | | | | | | | | | | | | | ☐ Yes ☐ No | | | | | | | | | | | | | | | | | | | | | | |
| 32. Fainting / blackouts | | | | | | | | | | | | | | | | | | | | | | | | | | | | | | | | | ☐ Yes ☐ No | | | | | | | | | | | | | | | | | | | | | | |
| **Gastro-intestinal / Genitourinary** | | | | | | | | | | | | | | | | | | | | | | | | | | | | | | | | | | | | | | | | | | | | | | | | | | | | | | | |
| 33. Diarrhoea | | | | | | | | | | | | | | | | | | | | | | | | | | | | | | | | | ☐ Yes ☐ No | | | | | | | | | | | | | | | | | | | | | | |
| 34. Constipation | | | | | | | | | | | | | | | | | | | | | | | | | | | | | | | | | ☐ Yes ☐ No | | | | | | | | | | | | | | | | | | | | | | |
| 35. Nausea/vomiting | | | | | | | | | | | | | | | | | | | | | | | | | | | | | | | | | ☐ Yes ☐ No | | | | | | | | | | | | | | | | | | | | | | |
| 36. Abdominal pain | | | | | | | | | | | | | | | | | | | | | | | | | | | | | | | | | ☐ Yes ☐ No | | | | | | | | | | | | | | | | | | | | | | |
| 37. Loss of appetite | | | | | | | | | | | | | | | | | | | | | | | | | | | | | | | | | ☐ Yes ☐ No | | | | | | | | | | | | | | | | | | | | | | |
| 38. Loss of control of passing urine | | | | | | | | | | | | | | | | | | | | | | | | | | | | | | | | | ☐ Yes ☐ No | | | | | | | | | | | | | | | | | | | | | | |
| 39. Loss of control of opening your bowels | | | | | | | | | | | | | | | | | | | | | | | | | | | | | | | | | ☐ Yes ☐ No | | | | | | | | | | | | | | | | | | | | | | |
| 40. Weight loss | | | | | | | | | | | | | | | | | | | | | | | | | | | | | | | | | ☐ Yes ☐ No | | | | | | | | | | | | | | | | | | | | | | |
| 41. Stomach pain | | | | | | | | | | | | | | | | | | | | | | | | | | | | | | | | | ☐ Yes ☐ No | | | | | | | | | | | | | | | | | | | | | | |
| 42. Erectile Dysfunction | | | | | | | | | | | | | | | | | | | | | | | | | | | | | | | | | ☐ Yes ☐ No ☐ N/A | | | | | | | | | | | | | | | | | | | | | | |
| **Skin** | | | | | | | | | | | | | | | | | | | | | | | | | | | | | | | | | | | | | | | | | | | | | | | | | | | | | | | |
| 43. Skin rash | | | | | | | | | | | | | | | | | | | | | | | | | | | | | | | | | ☐ Yes ☐ No | | | | | | | | | | | | | | | | | | | | | | |
| **If yes, please tick all body areas that apply:**  **☐**Face ☐Trunk (stomach or back) ☐Arms ☐Legs ☐Buttocks ☐Toes ☐Fingers | | | | | | | | | | | | | | | | | | | | | | | | | | | | | | | | | | | | | | | | | | | | | | | | | | | | | | | |
| 44. Lumpy lesions (purple/pink/bluish) on toes | | | | | | | | | | | | | | | | | | | | | | | | | | | | | | | | | ☐ Yes ☐ No | | | | | | | | | | | | | | | | | | | | | | |
| 45. Have you noticed any hair loss? | | | | | | | | | | | | | | | | | | | | | | | | | | | | | | | | | ☐ Yes ☐ No | | | | | | | | | | | | | | | | | | | | | | |
| 46. Bleeding | | | | | | | | | | | | | | | | | | | | | | | | | | | | | | | | | ☐ Yes ☐ No | | | | | | | | | | | | | | | | | | | | | | |
| If yes, please specify bleeding site: ________________________________________________________ | | | | | | | | | | | | | | | | | | | | | | | | | | | | | | | | | | | | | | | | | | | | | | | | | | | | | | | |
| 47. Any other ongoing symptoms? | | | | | | | | | | | | | | | | | | | | | | | | | | | | | | | | | **☐** Yes ☐ No | | | | | | | | | | | | | | | | | | | | | | |
| Please specify: ________________________________________________________ | | | | | | | | | | | | | | | | | | | | | | | | | | | | | | | | | | | | | | | | | | | | | | | | | | | | | | | |
| i) Are you or your family concerned that you have lost significant weight (5-10%) in the past 12 months, or may now be underweight? | | | | | | | | | | | | | | | | | | | | | | | | | | | | | | | | | | | | | | | | | | | | | | | | | Yes ☐ No **☐** | | | | | | |
| j) Regarding your appetite or interest in eating (since COVID-19), please rank your appetite or interest in eating on a scale of 0-5 (0 being same as usual/no problems, 5 being very severe problems/reduction) | | | | | | | | | | | | | | | | | | | | | | | | | | | | | | | | | ☐0 ☐1 ☐2 ☐3 ☐4 ☐5 | | | | | | | | | | | | | | | | | | | | | | |
| k) Were you admitted to Intensive Care (ITU) during admission? | | | | | | | | | | | | | | | | | | | | | | | | | | | | | | | | | **☐** Yes ☐ No  *(if no, please skip to section I on the next page)* | | | | | | | | | | | | | | | | | | | | | | |
| 1. Laryngeal/ airway complications   Have you developed any changes in the sensitivity of your throat such as troublesome cough or noisy breathing?  If Yes: rate the significance of impact on a scale of 0-5 (0 being no impact, 5 being significant impact) | | | | | | | | | | | | | | | | | | | | | | | | | | | | | | | | | ☐ Yes ☐ No | | | | | | | | | | | | | | | | | | | | | | |
|  |  |  |  |  |  |  |  |  |  |  |  |  |  |  |  |  |  |  |  |  |  |  |  |  |  |  |  |  |  |  |  |  | ☐0 ☐1 ☐2 ☐3 ☐4 ☐5 | | | | | | | | | | | | | | | | | | | | | | |
| 2. Swallowing   Are you having difficulties eating, drinking or swallowing such as coughing, choking or avoiding any food or drinks?  If Yes: rate the significance of impact on a scale of 0-5 (0 being no impact, 5 being significant impact) | | | | | | | | | | | | | | | | | | | | | | | | | | | | | | | | | ☐ Yes ☐ No | | | | | | | | | | | | | | | | | | | | | | |
|  |  |  |  |  |  |  |  |  |  |  |  |  |  |  |  |  |  |  |  |  |  |  |  |  |  |  |  |  |  |  |  |  | ☐0 ☐1 ☐2 ☐3 ☐4 ☐5 | | | | | | | | | | | | | | | | | | | | | | |
| 3. Voice   Have you or your family noticed any changes to your voice such as difficulty being heard, altered quality of the voice, your voice tiring by the end of the day or an inability to alter the pitch of your voice?  If Yes: rate the significance of impact on a scale of 0-5 (0 being no impact, 5 being significant impact) | | | | | | | | | | | | | | | | | | | | | | | | | | | | | | | | | ☐ Yes ☐ No | | | | | | | | | | | | | | | | | | | | | | |
|  |  |  |  |  |  |  |  |  |  |  |  |  |  |  |  |  |  |  |  |  |  |  |  |  |  |  |  |  |  |  |  |  | ☐0 ☐1 ☐2 ☐3 ☐4 ☐5 | | | | | | | | | | | | | | | | | | | | | | |
| l) The next questions ask about difficulties you may have doing certain activities because of a **HEALTH PROBLEM.** | | | | | | | | | | | | | | | | | | | | | | | | | | | | | | | | | | | | | | | | | | | | | | | | | | | | | | | |
|  | | | | | | | | | | | | | | | | | | | | | | | | | | | | **(Mark the correct answer with a tick in the box)** | | | | | | | | | | | | | | | | | | | | | | | | | | | |
| 1. Do you have difficulty seeing, even if wearing glasses? | | | | | | | | | | | | | | | | | | | | | | | | | | | | ☐ No – no difficulty | | | | | | | | | | | | | | | | | | | | | | | | | | | |
|  |  |  |  |  |  |  |  |  |  |  |  |  |  |  |  |  |  |  |  |  |  |  |  |  |  |  |  | ☐ Yes – some difficulty | | | | | | | | | | | | | | | | | | | | | | | | | | | |
|  |  |  |  |  |  |  |  |  |  |  |  |  |  |  |  |  |  |  |  |  |  |  |  |  |  |  |  | ☐ Yes – a lot of difficulty | | | | | | | | | | | | | | | | | | | | | | | | | | | |
|  |  |  |  |  |  |  |  |  |  |  |  |  |  |  |  |  |  |  |  |  |  |  |  |  |  |  |  | ☐ Cannot do at all | | | | | | | | | | | | | | | | | | | | | | | | | | | |
| 2. Do you have difficulty hearing, even if using a hearing aid? | | | | | | | | | | | | | | | | | | | | | | | | | | | | ☐ No – no difficulty | | | | | | | | | | | | | | | | | | | | | | | | | | | |
|  |  |  |  |  |  |  |  |  |  |  |  |  |  |  |  |  |  |  |  |  |  |  |  |  |  |  |  | ☐ Yes – some difficulty | | | | | | | | | | | | | | | | | | | | | | | | | | | |
|  |  |  |  |  |  |  |  |  |  |  |  |  |  |  |  |  |  |  |  |  |  |  |  |  |  |  |  | ☐ Yes – a lot of difficulty | | | | | | | | | | | | | | | | | | | | | | | | | | | |
|  |  |  |  |  |  |  |  |  |  |  |  |  |  |  |  |  |  |  |  |  |  |  |  |  |  |  |  | ☐ Cannot do at all | | | | | | | | | | | | | | | | | | | | | | | | | | | |
| 3. Do you have difficulty walking or climbing steps? | | | | | | | | | | | | | | | | | | | | | | | | | | | | ☐ No – no difficulty | | | | | | | | | | | | | | | | | | | | | | | | | | | |
|  |  |  |  |  |  |  |  |  |  |  |  |  |  |  |  |  |  |  |  |  |  |  |  |  |  |  |  | ☐ Yes – some difficulty | | | | | | | | | | | | | | | | | | | | | | | | | | | |
|  |  |  |  |  |  |  |  |  |  |  |  |  |  |  |  |  |  |  |  |  |  |  |  |  |  |  |  | ☐ Yes – a lot of difficulty | | | | | | | | | | | | | | | | | | | | | | | | | | | |
|  |  |  |  |  |  |  |  |  |  |  |  |  |  |  |  |  |  |  |  |  |  |  |  |  |  |  |  | ☐ Cannot do at all | | | | | | | | | | | | | | | | | | | | | | | | | | | |
| 4. Do you have difficulty remembering or concentrating? | | | | | | | | | | | | | | | | | | | | | | | | | | | | ☐ No – no difficulty | | | | | | | | | | | | | | | | | | | | | | | | | | | |
|  |  |  |  |  |  |  |  |  |  |  |  |  |  |  |  |  |  |  |  |  |  |  |  |  |  |  |  | ☐ Yes – some difficulty | | | | | | | | | | | | | | | | | | | | | | | | | | | |
|  |  |  |  |  |  |  |  |  |  |  |  |  |  |  |  |  |  |  |  |  |  |  |  |  |  |  |  | ☐ Yes – a lot of difficulty | | | | | | | | | | | | | | | | | | | | | | | | | | | |
|  |  |  |  |  |  |  |  |  |  |  |  |  |  |  |  |  |  |  |  |  |  |  |  |  |  |  |  | ☐ Cannot do at all | | | | | | | | | | | | | | | | | | | | | | | | | | | |
| 5. Do you have difficulty (with self-care such as) washing all over or dressing? | | | | | | | | | | | | | | | | | | | | | | | | | | | | ☐ No – no difficulty | | | | | | | | | | | | | | | | | | | | | | | | | | | |
|  |  |  |  |  |  |  |  |  |  |  |  |  |  |  |  |  |  |  |  |  |  |  |  |  |  |  |  | ☐ Yes – some difficulty | | | | | | | | | | | | | | | | | | | | | | | | | | | |
|  |  |  |  |  |  |  |  |  |  |  |  |  |  |  |  |  |  |  |  |  |  |  |  |  |  |  |  | ☐ Yes – a lot of difficulty | | | | | | | | | | | | | | | | | | | | | | | | | | | |
|  |  |  |  |  |  |  |  |  |  |  |  |  |  |  |  |  |  |  |  |  |  |  |  |  |  |  |  | ☐ Cannot do at all | | | | | | | | | | | | | | | | | | | | | | | | | | | |
| 6. Using your usual (customary) language, do you have difficulty communicating, for example understanding or being understood? | | | | | | | | | | | | | | | | | | | | | | | | | | | | ☐ No – no difficulty | | | | | | | | | | | | | | | | | | | | | | | | | | | |
|  |  |  |  |  |  |  |  |  |  |  |  |  |  |  |  |  |  |  |  |  |  |  |  |  |  |  |  | ☐ Yes – some difficulty | | | | | | | | | | | | | | | | | | | | | | | | | | | |
|  |  |  |  |  |  |  |  |  |  |  |  |  |  |  |  |  |  |  |  |  |  |  |  |  |  |  |  | ☐ Yes – a lot of difficulty | | | | | | | | | | | | | | | | | | | | | | | | | | | |
|  |  |  |  |  |  |  |  |  |  |  |  |  |  |  |  |  |  |  |  |  |  |  |  |  |  |  |  | ☐ Cannot do at all | | | | | | | | | | | | | | | | | | | | | | | | | | | |
| m) How many units of alcohol, on average, do you consume per week? (<https://www.drinkaware.co.uk/tools/unit-and-calorie-calculator>) | | | | | | | | | | | | | | | | | | | | | | | | | | | | | | | | | | | | | | | | | | | | | | | | | | **☐ ☐ ☐** units/week | | | | | |
| n) Do you or have you ever smoked cigarettes? | | | | | | | | | | | | | | | | | | | | | | | | | | | | | | | Never | | | | | | | | | | | | | | | | | | | | **☐** | | | | |
|  | | | | | | | | | | | | | | | | | | | | | | | | | | | | | | | Ex-smoker | | | | | | | | | | | | | | | | | | | | **☐** | | | | |
|  | | | | | | | | | | | | | | | | | | | | | | | | | | | | | | | Current smoker | | | | | | | | | | | | | | | | | | | | **☐** | | | | |
| o) Do you currently use an e-cigarette or vape? | | | | | | | | | | | | | | | | | | | | | | | | | | | | | | | | | | | | | Yes ☐ No **☐** | | | | | | | | | | | | | | | | | | |
| p) Have you made lifestyle changes since your COVID-19 infection?   (mark the correct answer with a tick in the box) | | | | | | | | | | | | | | | | | | | | | | | | | | | | | | | | | | | | | | | | | | | | | | | | | | | | | | | |
|  | | | | | | | | | **I do this more often** | | | | | | | | | | | | | | | **I do this less often** | | | | | | | | | | | | | | | | | | | | **No difference** | | | | | | | | | | **N/A** | |
| **Smoking** | | | | | | | | | **☐** | | | | | | | | | | | | | | | **☐** | | | | | | | | | | | | | | | | | | | | **☐** | | | | | | | | | | **☐** | |
| **Drinking alcohol** | | | | | | | | | **☐** | | | | | | | | | | | | | | | **☐** | | | | | | | | | | | | | | | | | | | | **☐** | | | | | | | | | | **☐** | |
| **Eating healthy food** | | | | | | | | | **☐** | | | | | | | | | | | | | | | **☐** | | | | | | | | | | | | | | | | | | | | **☐** | | | | | | | | | | **☐** | |
| **Physical activity** (include walking, cycling, & other activities) | | | | | | | | | **☐** | | | | | | | | | | | | | | | **☐** | | | | | | | | | | | | | | | | | | | | **☐** | | | | | | | | | | **☐** | |
| q) Please describe your occupation / working status **today**?   (If you are currently furloughed please select your status prior to furlough) | | | | | | | | | | | | | | | | | | | | | | | | | | | | | | | | | | | | | | | | | | | | | | | | | | | | | | | |
| ☐ Working Full-time | | | | | | | | | | | | | | | | | | | | | | | | | | | | | | | | | | | | | | | | | ☐ Working Part-time | | | | | | | | | | | | | | |
| ☐ Full time carer (children or other) | | | | | | | | | | | | | | | | | | | | | | | | | | | | | | | | | | | | | | | | | ☐ Unemployed | | | | | | | | | | | | | | |
| ☐ Unable to work due to chronic illness | | | | | | | | | | | | | | | | | | | | | | | | | | | | | | | | | | | | | | | | | ☐ Student | | | | | | | | | | | | | | |
| ☐ Retired | | | | | | | | | | | | | | | | | | | | | | | | | | | | | | | | | | | | | | | | | ☐ Medically retired | | | | | | | | | | | | | | |
| ☐ Prefer not to say | | | | | | | | | | | | | | | | | | | | | | | | | | | | | | | | | | | | | | | | |  | | | | | | | | | | | | | | |
| **Compared to before your COVID-19 illness** is your main occupation/working status: | | | | | | | | | | | | | | | | | | | | | | | | | | | | | | | | | | | | | | | | | | | | | | | | | | | | | | | |
| ☐ Same as before | | | | | | | | | | ☐ Different from before | | | | | | | | | | | | | | | | | | | | | | | | | | ☐ Prefer not to say | | | | | | | | | | | | | | | | | | | |
| If different, why did your occupation/working status change? | | | | | | | | | | | | | | | | | | | | | | | | | | | | | | | | | | | | | | | | | | | | | | | | | | | | | | | |
| ☐ Poor health | | | | | | | | | | | | | | | | | | | | | | | | | | | | | | | ☐ New caring responsibility | | | | | | | | | | | | | | | | | | | | | | | | |
| ☐ Working hours reduced by employer | | | | | | | | | | | | | | | | | | | | | | | | | | | | | | | ☐ Made redundant | | | | | | | | | | | | | | | | | | | | | | | | |
| ☐ Sick leave | | | | | | | | | | | | | | | | | | | | | | | | | | | | | | | ☐ Prefer not to say | | | | | | | | | | | | | | | | | | | | | | | | |
| ☐ Other (please specify): _______________________________________________ | | | | | | | | | | | | | | | | | | | | | | | | | | | | | | | | | | | | | | | | | | | | | | | | | | | | | | | |
| Have you ever been furloughed? | | | | | | | | | | | | | | | | | | | | | | | | | | | | | | | | | | | | | |  | | | | | | | | | | | | | | | | | |
| Yes | | | | | | | ☐ | | | | | | | | | | | No | | | | | | | **☐** | | | | | | | | | | | | | | | | | | | | | | | | | | | | | | |
| If yes, when were you furloughed? (select all that apply) | | | | | | | | | | | | | | | | | | | | | | | | | | | | | | | | | | | | | | | | | | | | | | | | | | | | | | | |
| Before you were admitted to hospital with COVID-19 | | | | | | | ☐ | | | | | | | | | | Since you were discharged from hospital after COVID-19 | | | | | | | | | | | | | | | | | | | | | | | | | | | | | | | ☐ | | | | | | | |
| **If yes, for how long have you been/ were you furloughed in total? ☐ ☐** months (to the nearest month)  (If furloughed for less than 1 month please enter 1. If you were on flexi-furlough or intermittently furloughed please estimate the total number of months you have been/ were furloughed for.) | | | | | | | | | | | | | | | | | | | | | | | | | | | | | | | | | | | | | | | | | | | | | | | | | | | | | | | |
| **Has your illness affected your ability to do your usual work?** | | | | | | | | | | | | | | | | | | | | | | | | | | | | | | | | Yes ☐ No **☐** | | | | | | | | | | | | | | | | | | | | | | | |
| r) Regarding loneliness and mental health, please complete the following (for each statement select one answer): | | | | | | | | | | | | | | | | | | | | | | | | | | | | | | | | | | | | | | | | | | | | | | | | | | | | | | | |
|  | | | | | | | | | | | | | | | | | | | | | | | **Hardly ever** | | | | | | | | | | | | | | | | **Some of the time** | | | | | | | | | | | | | | **Often** | | |
| How often do you feel that you lack companionship? | | | | | | | | | | | | | | | | | | | | | | | **☐** | | | | | | | | | | | | | | | | **☐** | | | | | | | | | | | | | | **☐** | | |
| How often do you feel left out? | | | | | | | | | | | | | | | | | | | | | | | **☐** | | | | | | | | | | | | | | | | **☐** | | | | | | | | | | | | | | **☐** | | |
| How often do you feel isolated from others? | | | | | | | | | | | | | | | | | | | | | | | **☐** | | | | | | | | | | | | | | | | **☐** | | | | | | | | | | | | | | **☐** | | |
| How often do you feel lonely? | | | | | | | | | | | | | | | | | | | | | | | **☐** | | | | | | | | | | | | | | | | **☐** | | | | | | | | | | | | | | **☐** | | |
| s) Overall, how satisfied are you with your life nowadays, where 0 means 'not at all' and 10 means 'completely'? | | | | | | | | | | | | | | | | | | | | | | | | | | | | | | | | | | | | | | | | | | | | | | | | | | | | | | | |
| 0 | 1 | | | 2 | | 3 | | | | | | | 4 | | | 5 | | | | | 6 | | | | | | | | | 7 | | | 8 | | | | | | | | | | | | 9 | | | | | 10 | | | | | |
| **☐** | **☐** | | | **☐** | | **☐** | | | | | | | **☐** | | | **☐** | | | | | **☐** | | | | | | | | | **☐** | | | **☐** | | | | | | | | | | | | **☐** | | | | | **☐** | | | | | |
| Not at all satisfied |  | | | | | | | | | | | | | | | | | | | | | | | | | | | | | | | | | | | | | | | | | | | | | | | | | Completely satisified | | | | | |
| t) Have you felt feverish recently? | | | | | | | | | | | | | | | | | | | | | | | | | | | | | ☐ Yes | | | | | | | | | | | | | ☐ No | | | | | | | | ☐ Not sure | | | | | |
| If yes, roughly when did you last feel feverish? | | | | | | | | | | | | | | | | | | | | | | | | | | | | | | | | | | | | | | | | | | | | | | | | | | | | | | | |
| ☐ within the last 7 days | | | | | | | | | | | | ☐ between 1 to 2 weeks ago | | | | | | | | | | | | | | | | | | | | | | | ☐ between 2 to 4 weeks ago | | | | | | | | | | | | | | | | | | | | |
| ☐ between 1 to 2 months ago | | | | | | | | | | | | ☐ between 2 to 3 months ago | | | | | | | | | | | | | | | | | | | | | | | | | | | | |  | | | | | | | | | | | | | | |
| If yes, what was the cause of your most recent feverish illness? | | | | | | | | | | | | | | | | | | | | | | | | | | | | | | | | | | | | | | | | | | | | | | | | | | | | | | | |
| ☐ COVID-19 | | ☐ | Other respiratory infection (cough/cold/sore throat) | | | | | | | | | | | | | | | | | ☐ | | | | Stomach infection (diarrhoea/vomiting) | | | | | | | | | | | | | | | | | | | | | | | ☐ Urinary infection | | | | | | | | |
| ☐ TB | | ☐ Other: Specify _____________ | | | | | | | | | | | | | | | | | | ☐ Unknown | | | | | | | | | | | | | | | | | | | | | | | | | | | ☐ Prefer not to say | | | | | | | | |
| u) Regarding hearing and balance disturbance: | | | | | | | | | | | | | | | | | | | | | | | | | | | | | | | | | | | | | | | | | | | | | | | | | | | | | | | |

| Have you had noises (such as ringing or buzzing) in your head or in one or both ears that lasts for more than 5 minutes at a time? | **☐** Yes, most or all of the time  **☐** Yes, a lot of the time  **☐** Yes, some of the time  **☐** No, not in the past year  **☐** No, never  **☐** Do not know/Prefer not to answer | |
| --- | --- | --- |
| Have you ever suffered from? |  |  |
| a) Attacks of dizziness in which things seem to spin around you? | **☐ Yes** | **☐ No** |
| b) Attacks of dizziness in which you seem to move? | **☐ Yes** | **☐ No** |

| v) **Vaccination** | | |
| --- | --- | --- |
| Have you received a SARS-CoV-2 (Coronavirus) vaccine first dose? | Yes **☐** No **☐** N/K **☐**  If yes, on what date? _ _ / _ _ _ / _ _ _ _  Or date N/K **☐** | |
| If yes, do you know which vaccine you received? | Yes **☐** No **☐** | |
|  | Oxford/Astra-Zenca | **☐** |
|  | Pfizer/ Bio-N-Tec | **☐** |
|  | Moderna | **☐** |
|  | Other | **☐** Specify:_________________ |
| Have you received a SARS-CoV-2 (Coronavirus) vaccine second dose? | Yes **☐** No **☐** N/K **☐**  If yes, on what date? _ _ / _ _ _ / _ _ _ _  Or date N/K **☐** | |
| If yes, do you know which vaccine you received? | Yes **☐** No **☐** | |
|  | Oxford/Astra-Zenca | **☐** |
|  | Pfizer/ Bio-N-Tec | **☐** |
|  | Moderna | **☐** |
|  | Other | **☐** Specify:_________________ |
